# A Comparison of Diagnostic Models and Prognostic Scores of ACLF: Towards Global Harmonization

**DOI:** 10.64898/2026.04.29.26352045

**Authors:** Meiqian Hu, Jinjin Luo, Nipun Verma, Pratibha Garg, Sunil Taneja, Juan Antonio Carbonell-Asins, María Pilar Ballester, Tingting Qi, Sina Jameie-Oskooei, Qun Cai, Xi Liang, Jiaqi Li, Tianzhou Wu, Jiang Li, Peng Li, Qian Zhou, Jiaojiao Xin, Dongyan Shi, Jing Jiang, Wei Qiang, Changze Hong, Xin Chen, Bing Zhu, Tingting Feng, Jianming Zheng, Yuxian Huang, Feng Ye, Bingliang Lin, Jinjun Chen, Rajeshwar P Mookerjee, Yan Huang, Shaoli You, Cornelius Engelmann, Yu Chen, Ajay Duseja, Jun Li, Rajiv Jalan

## Abstract

**Background and Aims:** Acute-on-chronic liver failure (ACLF) is associated with high short-term mortality, but substantial heterogeneity among existing diagnostic and prognostic models results in inconsistent patient identification and risk assessment. We conducted a systematic head-to-head comparison of major ACLF diagnostic and prognostic models to evaluate concordance, short-term mortality prediction and clinical utility, with the goal of informing harmonization of ACLF assessment.

**Methods:** We analysed 3,370 patients with acute decompensation of cirrhosis in the COSSH cohort, with external validation in an independent Ambi-Spective cohort from India (n=2,055). Five ACLF diagnostic models were evaluated for identification of patients at risk of 28-day mortality. Reclassification was assessed using net reclassification improvement. Prognostic scores were compared using concordance index, integrated discrimination improvement, calibration, and decision-curve analysis.

**Results:** Diagnostic frameworks identified markedly different proportions of ACLF. A-TANGO and COSSH-ACLF classified the largest high-risk populations while maintaining substantial short-term mortality and balanced sensitivity–specificity profiles. Compared with COSSH-ACLF, A-TANGO improved net reclassification by 7.7%, with further gains versus EASL-CLIF (11.8%), APASL-ACLF (36.4%), and NACSELD-ACLF (45.9%). In the external cohort, A-TANGO and COSSH-ACLF showed similar discrimination and identified comparable proportions of patients. Combined application of the two models delineated three clinically meaningful strata, identifying a discordant intermediate-risk group with approximately 11% 28–day mortality. Among prognostic scores, COSSH-ACLF II and A-TANGO OF scores demonstrated strong and complementary performance across cohorts.

**Conclusions:** Outcome-anchored ACLF definitions converge in identifying patients at highest short-term risk across diverse populations. Alignment between A-TANGO and COSSH-ACLF, together with identification of an intermediate-risk phenotype, supports a data-driven framework for improving consistency and advancing global harmonization of ACLF diagnosis and risk stratification.

## Introduction

Acute-on-chronic liver failure (ACLF) is a life-threatening syndrome characterized by acute decompensation, organ failure, and high short-term mortality.^1–4^ Accurate identification and risk stratification are critical for timely management, including intensive care and consideration of liver transplantation (LT).^5–7^ Several major ACLF diagnostic models are widely used worldwide, but the European Association for the Study of the Liver-Chronic Liver Failure Consortium (EASL-CLIF)^1,8^, Chinese Group on the Study of Severe Hepatitis B (COSSH)-ACLF^9–11^, North American Consortium for the Study of End-Stage Liver Disease (NACSELD)-ACLF^12^, and Asian Pacific Association for the Study of the Liver (APASL)-ACLF.^13,14^ Recently, the EASL-CLIF model was modified to develop and validate the A-TANGO diagnostic model to enhance patient identification and risk stratification. The A-TANGO model demonstrated more favorable performance compared with the EASL-CLIF diagnostic model in global cohorts.^15^ However, its performance relative to other established diagnostic models remains unclear. Likewise, the COSSH diagnostic model has also been reported to outperform the EASL-CLIF diagnostic model^10^ but direct comparisons across multiple diagnostic models are limited. Despite widespread adoption, contemporary ACLF diagnostic models remain fragmented across regions, leading to inconsistent identification of ACLF and variable estimates of short-term risk within the same population. Such discrepancies may directly influence clinical decision-making, including intensive care allocation, LT prioritization, and enrolment in clinical trials. This lack of alignment complicates clinical decision-making and underscores the need for greater harmonization in ACLF evaluation.

Multiple prognostic scores have been developed for patients with ACLF, including the Chronic Liver Failure Consortium (CLIF-C) ACLF score, CLIF-C Organ Failure (OF) score, COSSH-ACLF, COSSH-ACLF II, and the more recent A-TANGO scores. In addition, the score for End-Stage Liver Disease-Sodium (MELD-Na)^16^, though not specifically designed for ACLF, is widely used in clinical practice. However, a systematic, head-to-head evaluation of these prognostic scores is currently lacking, both within populations defined by different ACLF diagnostic models and across the broader spectrum of patients hospitalized with acute decompensation of cirrhosis. Furthermore, the reliance of some ACLF scores on dynamic parameters that can be influenced by medical interventions (e.g., white blood cell count (WBC) or C-reactive protein (CRP))^17^ or on organ failure definitions (e.g., hepatic encephalopathy (HE))^18^ that are difficult to standardize consistently may affect their generalizability.

To address these gaps, we performed a comprehensive head-to-head evaluation of major ACLF diagnostic models and prognostic scores in a large multicenter cohort of patients with acute decompensation of cirrhosis, with external validation in two independent populations. Anchored by the COSSH cohort from China and the Ambi-Spective cohort from India, our study aimed to delineate alignment and discordance in patient identification, risk stratification and short-term mortality prediction, thereby informing ongoing efforts toward harmonization of ACLF evaluation.

## Methods

### Study design

This study was conducted in a large multicenter cohort of hospitalized patients with acute decompensation of cirrhosis from the COSSH database, with external validation performed in an independent Ambi-Spective cohort from India. We systematically evaluated major ACLF diagnostic models and prognostic scores to examine their performance in patient identification, risk stratification, and short-term mortality prediction. Five widely used ACLF diagnostic models were compared, and ten prognostic scores were assessed using complementary statistical approaches, including discrimination, reclassification, calibration and clinical utility analyses.

### Patients

The primary population was derived from the multicentre COSSH cohort, including consecutive hospitalized patients with acute decompensation of cirrhosis of all etiologies between January 2018 and August 2023. Eligible patients presented with typical complications (ascites, hepatic encephalopathy, gastrointestinal bleeding, or infection). Exclusion criteria included age <18 or >80 years, pregnancy, malignancy, severe extrahepatic organ dysfunction unrelated to liver disease, immunosuppressive or psychotropic therapy, or human immunodeficiency virus infection. A total of 3,370 patients were included, of whom 199 and 276 underwent liver transplantation within 28 and 90 days. External validation was performed in an independent Ambi-Spective cohort from India, comprising 2,055 hospitalized patients with decompensated cirrhosis between 2015 and 2023. Patients were followed for 90 days, and none underwent liver transplantation. Those with missing key baseline variables were excluded. The study protocols were approved by the respective institutional ethics committees (COSSH: No.2017-51 and No.2022-995; India: PGI/IEC/2021/001346). Written informed consent was obtained from prospectively recruited participants, with a waiver granted for retrospective data where applicable. All procedures conformed to the Declaration of Helsinki and the Declaration of Istanbul.

### Diagnostic models and prognostic scores

ACLF was diagnosed at admission using five diagnostic models: A-TANGO, EASL-CLIF, COSSH-ACLF, APASL-ACLF, and NACSELD-ACLF. For prognostic assessment, a total of ten scores were calculated, comprising the A-TANGO ACLF-CRP, A-TANGO ACLF-WBC, COSSH-ACLF, COSSH-ACLF II, CLIF-C ACLF, the APASL ACLF Research Consortium (AARC), NACSELD-ACLF, A-TANGO OF, CLIF-OF, and MELD-Na scores. Due to missing serum lactate measurements, AARC scores were only available for only 388 patients. Detailed descriptions of the five diagnostic models and the calculation formulas for the ten prognostic scores are provided in Supplemental Tables 1 and 2.

### Statistics

Continuous variables were presented as mean ± standard deviation for normally distributed data or median (interquartile range (Q1, Q3)) for non-normally distributed data, whereas categorical variables were summarized as counts and percentages. Normality was assessed using histograms, Q-Q plots, and the Kolmogorov–Smirnov test, and homogeneity of variance was evaluated using Levene’s test. For group comparisons of continuous variables, Student’s t-test (two groups) and one-way analysis of variance (ANOVA; multiple groups) were applied when both normality and homogeneity assumptions were satisfied. When variances were unequal, Welch’s t-test or Welch ANOVA was used. When either normality or variance assumptions were violated, the Mann–Whitney U test (two groups) or Kruskal–Wallis test (multiple groups) was applied. Categorical variables were compared using the χ² test or Fisher’s exact test when the expected frequency in any cell was <5. Diagnostic agreement between ACLF diagnostic models was assessed using McNemar’s test, with corresponding p values used to determine the significance of inter-model classification differences. Diagnostic performance metrics, including sensitivity, specificity, positive predictive value (PPV), and negative predictive value (NPV), were calculated using 2×2 contingency tables with 28–day LT-free mortality as the binary outcome. Kaplan–Meier survival analysis with the log-rank test was performed to compare survival outcomes across different ACLF grades. Liver transplantation was treated as a censoring event. The Net Reclassification Improvement (NRI)^19^ was calculated to assess differences in risk classification between diagnostic models. NRI was calculated based on ACLF grade reclassification defined by different diagnostic models, using 28–day LT-free mortality as the clinical outcome. Discriminatory ability was assessed by the concordance index (C-index), with C-indexes compared using a U-statistics-based estimator with a z-score test. And the Integrated Discrimination Improvement (IDI)^20^ was used to evaluate the overall improvement in predictive performance for predicting 28–day LT-free mortality. Calibration curves^21^ were constructed to assess the agreement between predicted and observed 28–day LT-free mortality. Decision curve analysis (DCA)^22^ was performed to evaluate the clinical net benefit of different ACLF prognostic scores across a range of threshold probabilities. Univariable and multivariable Cox proportional hazards regression analyses were performed to identify predictors of 90–day liver transplant-free mortality within the medium-risk group. In Cox models, liver transplantation was treated as a censoring event. Sensitivity analyses were performed to assess the robustness of the results. Missing data were handled using MICE with five imputations^23^, and results were pooled using Rubin’s rules. Variables with <5% missingness were analyzed using complete cases. For repeated measurements, mean baseline values were used. Bootstrap confidence intervals were derived with resampling applied as an outer loop around multiple imputation.^24^ All statistical analyses were conducted using R software (version 4.0.2) and SPSS software (version 25.0). A two-tailed P value <0.05 was considered statistically significant, and all P values are reported to two significant digits.

## Results

### Diagnostic concordance and risk stratification across five ACLF diagnostic models

A total of 3,370 patients with acute decompensated cirrhosis from the COSSH cohort were included. The five ACLF diagnostic models identified markedly different proportions of ACLF: COSSH-ACLF (n=1,155), A-TANGO (n=1,134), EASL-CLIF (n=706), APASL-ACLF (n=437), and NACSELD-ACLF (n=93). COSSH-ACLF and A-TANGO diagnostic models classified the largest number of patients as ACLF, followed by the EASL-CLIF and APASL-ACLF models, whereas the NACSELD-ACLF model identified only a small subset. To further characterize the concordance between models and the associated clinical risk, pairwise overlap is summarized in Figure 1A. A-TANGO overlapped most extensively with COSSH-ACLF (n=947), and this shared subgroup exhibited high 28- and 90-day LT-free mortality (36.8% and 54.8%).

**Figure 1.**
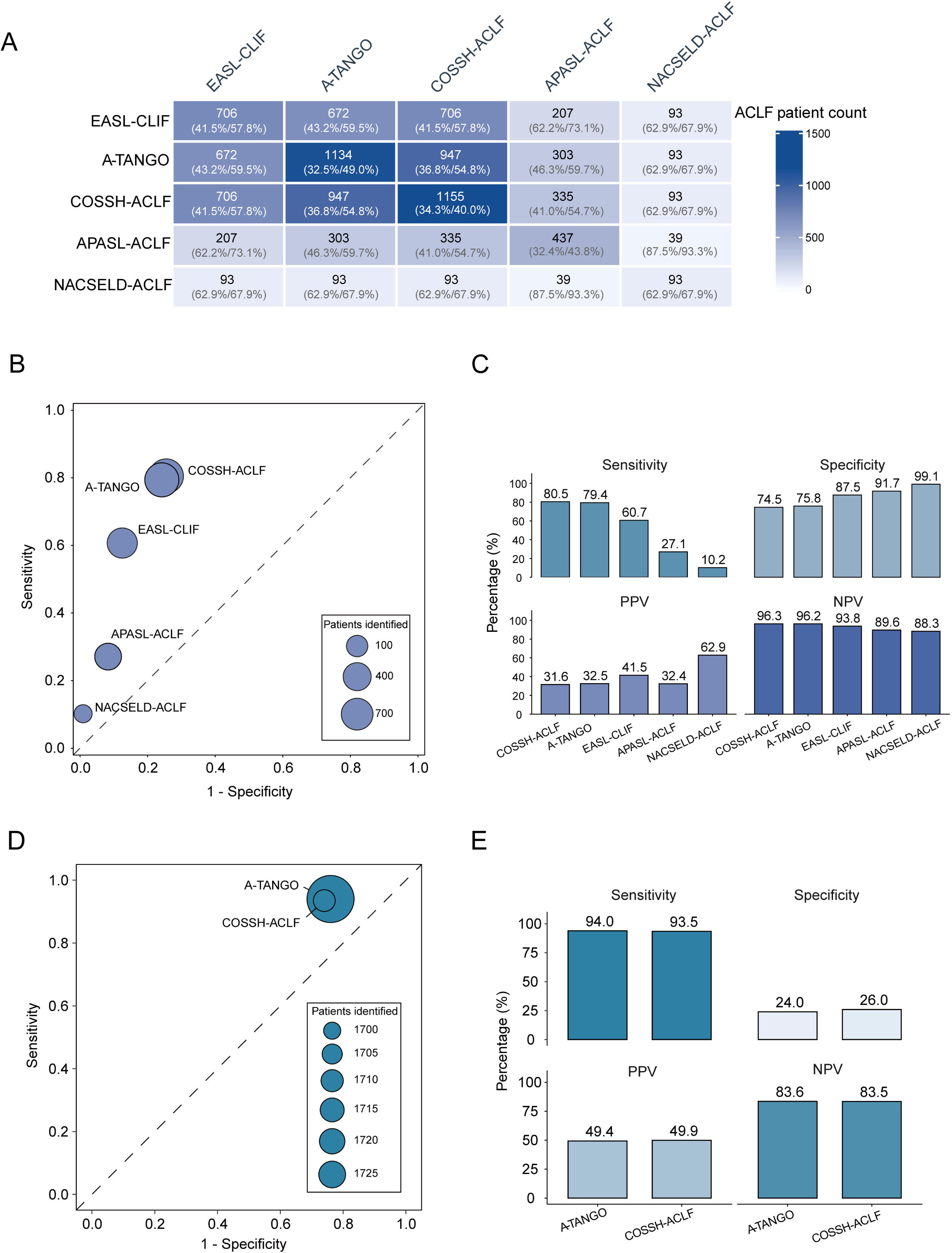
Evaluation of five ACLF diagnostic models and their prognostic implications. (A) Overlap of diagnostic models in the COSSH cohort (n=3,370). Diagonal values indicate the number of patients identified by each model; parentheses show 28- and 90-day liver transplant-free mortality. (B) Discrimination for 28-day liver transplant-free mortality in the COSSH cohort. Models were treated as binary variables; bubble size reflects the number of patients classified as ACLF. (C) Diagnostic performance corresponding to panel B, including sensitivity, specificity, PPV, and NPV. (D) Discrimination of A-TANGO and COSSH-ACLF models in the Indian Ambi-Spective cohort. (E) Corresponding diagnostic performance metrics in the Indian Ambi-Spective cohort. ACLF, acute-on-chronic liver failure; APASL, Asian Pacific Association for the Study of the Liver; COSSH, Chinese Group on the Study of Severe Hepatitis B; EASL-CLIF, European Association for the Study of the Liver-Chronic Liver Failure Consortium; NACSELD, North American Consortium for the Study of End-Stage Liver Disease; NPV, negative predictive value; PPV, positive predictive value.

We next evaluated the ability of each diagnostic model to discriminate 28-day LT-free mortality (Figures 1B, C). For each model, ACLF status was dichotomized (ACLF vs. no ACLF) and assessed against 28–day mortality (death vs. survival). In ROC space, the COSSH-ACLF and A-TANGO models showed the greatest separation between survivors and non-survivors, locating closest to the upper-left corner, whereas the EASL-CLIF model demonstrated intermediate discrimination. The APASL-ACLF and NACSELD-ACLF models were highly specific but substantially less sensitive, missing a considerable proportion of patients who subsequently died. Consistent with these findings, pairwise McNemar analyses demonstrated significant differences in ACLF classification between most models, while no significant difference was observed between A-TANGO and COSSH-ACLF models (P=0.43), indicating broadly similar classification behavior and risk stratification patterns (Supplemental Table S3). Although A-TANGO and COSSH-ACLF models labeled a larger proportion of patients as ACLF, these populations still exhibited high short-term mortality, suggesting improved risk enrichment rather than over-classification.

### ACLF grade distribution and net reclassification improvement of the different diagnostic models

Kaplan–Meier curves for 28–day survival were generated according to ACLF grades defined by the five diagnostic models. The A-TANGO, EASL-CLIF, and COSSH-ACLF diagnostic models demonstrated clear and graded separation of 28–day survival across increasing ACLF grades (Supplemental Figure S1A). Compared with EASL-CLIF, the A-TANGO model reassigned many patients to different severity grades, and the mortality observed in each grade followed a clearer stepwise pattern, better reflecting true 28–day risk (Table 1). Among patients classified as no ACLF by the EASL-CLIF model, 16.6% (407/2445) were reclassified into higher-risk categories (A-TANGO ACLF grades 1-3). Within this reclassified group, the observed 28–day LT-free mortality demonstrated a clear stepwise increase, rising from 16.8% in A-TANGO grade 1 to 30.6% in grade 2. Conversely, within the EASL-CLIF ACLF grade 1 cohort, 16.5% (29/176) of patients were down-classified to no ACLF by the A-TANGO diagnostic model and exhibited a low mortality rate of 10.3%. Meanwhile, patients who were up-classified to A-TANGO grades 2-3 experienced markedly higher mortality, ranging from 40.9% to 100.0%. For those in EASL-CLIF ACLF grades 2 and 3, the A-TANGO model effectively redistributed a significant proportion of patients across different severity grades, with mortality rates consistently aligning with the reclassified grades (ranging from 34.6% to 82.8%). These reclassification patterns collectively resulted in a significant net reclassification improvement of 11.8% for the A-TANGO model, underscoring its enhanced ability to discriminate patient risk. Comparable improvements were also observed when comparing the A-TANGO model with COSSH-ACLF, APASL-ACLF, and NACSELD-ACLF, with NRI values of 7.7%, 36.4%, and 45.9%. These findings suggest that the A-TANGO model enables more effective risk stratification.

**Table 1.** Distribution of ACLF grades at baseline and net reclassification improvement of the A-TANGO diagnostic model compared with other diagnostic models for 28–day liver transplant–free mortality.

| A-TANGO diagnostic model vs. EASL-CLIF diagnostic model |  |  |  |  |  |  |
| --- | --- | --- | --- | --- | --- | --- |
| EASL-CLIF diagnostic model | A-TANGO diagnostic model (NRI=11.8%) |  |  |  |  |  |
|  | No ACLF | ACLF grade 1 | ACLF grade 2 | ACLF grade 3 | ACLF grade 4 | Total |
| No ACLF |  |  |  |  |  |  |
| Number of patients. n (%) | 2038 (83.4) | 357 (14.6) | 49 (2.0) | 1 (0.04) | NA | 2445 (81.3) |
| 28-day mortality. n (%) | 76 (3.7) | 60 (16.8) | 15 (30.6) | 0 (0.0) | NA | 151 (6.2) |
| ACLF grade 1 |  |  |  |  |  |  |
| Number of patients. n (%) | 29 (16.5) | 124 (70.5) | 22 (12.5) | 1 (0.6) | NA | 176 (5.9) |
| 28-day mortality. n (%) | 3 (10.3) | 28 (22.6) | 9 (40.9) | 1 (100.0) | NA | 41 (23.3) |
| ACLF grade 2 |  |  |  |  |  |  |
| Number of patients. n (%) | NA | 107 (35.1) | 164 (53.8) | 33 (10.8) | 1 (0.3) | 305 (10.1) |
| 28-day mortality. n (%) | NA | 37 (34.6) | 80 (36.6) | 20 (60.6) | 0 (0.0) | 137 (44.9) |
| ACLF grade 3 |  |  |  |  |  |  |
| Number of patients. n (%) | NA | NA | 17 (21.0) | 35 (43.2) | 29 (35.8) | 81 (2.7) |
| 28-day mortality. n (%) | NA | NA | 6 (35.3) | 25 (71.4) | 24 (82.8) | 55 (67.9) |
| Total | 2067 (68.7) | 588 (19.6) | 252 (8.4) | 70 (2.3) | 30 (1.0) | 3007 |
| A-TANGO diagnostic model vs. COSSH-ACLF diagnostic model |  |  |  |  |  |  |
| COSSH-ACLF diagnostic model | A-TANGO diagnostic model (NRI=7.7%) |  |  |  |  |  |
|  | No ACLF | ACLF grade 1 | ACLF grade 2 | ACLF grade 3 | ACLF grade 4 | Total |
| No ACLF |  |  |  |  |  |  |
| Number of patients. n (%) | 1871 (92.3) | 154 (7.6) | 3 (0.1) | NA | NA | 2028 (67.4) |
| 28-day mortality. n (%) | 58 (3.1) | 17 (11.0) | 0 (0.0) | NA | NA | 75 (3.7) |
| ACLF grade 1 |  |  |  |  |  |  |
| Number of patients. n (%) | 196 (33.1) | 327 (55.1) | 68 (11.5) | 2 (0.3) | NA | 593 (19.7) |
| 28-day mortality. n (%) | 21 (10.7) | 71 (21.7) | 24 (35.3) | 1 (0.5) | NA | 117 (19.7) |
| ACLF grade 2 |  |  |  |  |  |  |
| Number of patients. n (%) | NA | 107 (35.1) | 164 (53.8) | 33 (10.8) | 1 (0.3) | 305 (10.1) |
| 28-day mortality. n (%) | NA | 37 (34.6) | 80 (48.8) | 20 (60.6) | 0 (0.0) | 137 (44.9) |
| ACLF grade 3 |  |  |  |  |  |  |
| Number of patients. n (%) | NA | NA | 17 (21.0) | 35 (43.2) | 29 (35.8) | 81 (2.7) |
| 28-day mortality. n (%) | NA | NA | 6 (35.3) | 25 (71.4) | 24 (82.8) | 55 (67.9) |
| Total | 2067 (68.7) | 588 (19.6) | 252 (8.4) | 70 (2.3) | 30 (1.0) | 3007 |
| A-TANGO diagnostic model vs. APASL-ACLF diagnostic model |  |  |  |  |  |  |
| APASL-ACLF diagnostic model | A-TANGO diagnostic model (NRI=36.4%) |  |  |  |  |  |
|  | No ACLF |  | ACLF |  | Total |  |
| No ACLF |  |  |  |  |  |  |
| Number of patients. n (%) | 1947 (72.5) |  | 739 (27.5) |  | 2686 (89.3) |  |
| 28-day mortality. n (%) | 68 (3.5) |  | 212 (28.7) |  | 280 (10.4) |  |
| ACLF |  |  |  |  |  |  |
| Number of patients. n (%) | 120 (37.4) |  | 201 (62.6) |  | 321 (10.7) |  |
| 28-day mortality. n (%) | 11 (9.2) |  | 93 (46.3) |  | 104 (32.4) |  |
| Total | 2067 (68.7) |  | 940 (31.3) |  | 3007 |  |
| A-TANGO diagnostic model vs. NACSELD-ACLF diagnostic model |  |  |  |  |  |  |
| NACSELD-ACLF | A-TANGO diagnostic model (NRI=45.9%) |  |  |  |  |  |

| diagnostic model | No ACLF | ACLF | Total |
| --- | --- | --- | --- |
| No ACLF |  |  |  |
| Number of patients. n (%) | 2067 (70.2) | 878 (29.8) | 2945 (97.9) |
| 28-day mortality. n (%) | 79 (3.8) | 266 (30.3) | 345 (11.7) |
| ACLF |  |  |  |
| Number of patients. n (%) | NA | 62 (100.0) | 62 (2.1) |
| 28-day mortality. n (%) | NA | 39 (62.9) | 39 (62.9) |
| Total | 2067 (68.7) | 940 (31.3) | 3007 |

ACLF, acute-on-chronic liver failure; APASL, Asian Pacific Association for the Study of the Liver; COSSH, Chinese Group on the Study of Severe Hepatitis B; EASL-CLIF, European Association for the Study of the Liver-Chronic Liver Failure Consortium; NACSELD, North American Consortium for the Study of End-Stage Liver Disease; NRI, net reclassification improvement.

### Clinical profiles across five ACLF diagnostic models

Clinical profiles across the five ACLF diagnostic models are summarized in Table 2. Complication patterns showed both overlap and variation across models. HE was most prevalent among patients classified by the EASL-CLIF and NACSELD-ACLF models, whereas ascites predominated in APASL-ACLF patients. Gastrointestinal bleeding was more common among NACSELD-ACLF patients. APASL-ACLF patients were more frequently associated with HBV-related liver disease and less commonly with alcohol-related liver disease. Patients fulfilling the NACSELD-ACLF definition exhibited more pronounced renal dysfunction and systemic inflammatory features, including higher creatinine, serum urea, white blood cell, and neutrophil counts, accompanied by lower hemoglobin, hematocrit, and platelet levels. Clinical features were broadly comparable between patients identified by the A-TANGO and COSSH-ACLF models, supporting substantial biological overlap between the two models. NACSELD-ACLF patients also exhibited the highest short-term mortality, accompanied by consistently elevated prognostic scores. Among patients diagnosed with ACLF, short-term outcomes varied across diagnostic models, with 28-/90-day mortality of 31.6%/48.6% for COSSH-ACLF, 32.5%/49.0% for A-TANGO, 41.5%/57.8% for EASL-CLIF, 32.4%/43.8% for APASL-ACLF and 62.9%/67.9% for NACSELD-ACLF.

**Table 2.** Characteristics of ACLF patients under five diagnostic models.

| Characteristic | A-TANGO<br>(n=1134) | EASL-CLIF<br>(n=706) | COSSH-ACLF<br>(n=1155) | APASL-ACLF<br>(n=437) | NACSELD-ACLF<br>(n=93) | P value |
| --- | --- | --- | --- | --- | --- | --- |
| Age (years) | 52 ± 12 | 52 ± 12 | 52 ± 12 | 53 ± 11 | 54 ± 12 | 0.30 |
| Male (no.) | 76.1% (863) | 75.2% (531) | 77.1% (891) | 76.2% (333) | 76.3% (71) | 0.92 |
| Complication (no.) |  |  |  |  |  |  |
| Hepatic encephalopathy | 32.3% (366) | 48.9% (345) | 30.0% (347) | 35.9% (157) | 78.5% (73) | <0.001 |
| Gastrointestinal bleeding | 14.6% (164) | 13.3% (93) | 11.6% (133) | 10.8% (47) | 27.2% (25) | <0.001 |
| Ascites | 75.3% (854) | 77.6% (548) | 76.8% (887) | 91.5% (400) | 67.7% (63) | <0.001 |
| Infection | 66.9% (759) | 65.9% (465) | 68.8% (795) | 65.0% (284) | 62.4% (58) | 0.42 |
| Etiologies (no.) |  |  |  |  |  |  |
| HBV related | 58.6% (665) | 59.2% (418) | 58.5% (676) | 69.6% (304) | 62.4% (58) | <0.001 |
| Alcohol related | 27.1% (307) | 26.9% (190) | 28.2% (326) | 19.7% (86) | 25.8% (24) | 0.014 |
| Autoimmune related | 6.6% (75) | 6.2% (44) | 7.4% (86) | 5.0% (22) | 2.2% (2) | 0.18 |
| Others | 14.2% (161) | 14.3% (101) | 13.4% (155) | 11.4% (50) | 17.2% (16) | 0.50 |
| Laboratory data |  |  |  |  |  |  |
| ALT (U/L) | 71.0 (31.0, 194.3) | 74.0 (31.0, 201.3) | 83.0 (36.0, 223.0) | 99.0 (41.5, 282.0) | 99.0 (35.0, 320.5) | <0.001 |
| AST (U/L) | 102.0 (55.0, 202.4) | 105.0 (55.0, 206.0) | 115.9 (64.0, 215.3) | 110.0 (66.0, 221.0) | 129.0 (58.8, 302.5) | 0.0076 |
| ALP (U/L) | 127.3 (92.0, 165.0) | 125.0 (92.0, 164.0) | 133.0 (100.2, 170.0) | 129.0 (98.0, 158.0) | 105.0 (78.0, 140.0) | <0.001 |
| ALB (g/L) | 29.4 ± 5.1 | 29.5 ± 5.1 | 29.5 ± 5.0 | 29.6 ± 5.0 | 30.1 ± 5.7 | 0.81 |
| TB (μmol/L) | 349.8 (205.5, 457.9) | 332.9 (232.6, 453.7) | 338.0 (252.5, 444.3) | 313.7 (194.0, 426.1) | 303.3 (90.0, 436.9) | 0.016 |
| TG (mmol/L) | 1.0 (0.8, 1.4) | 1.0 (0.8, 1.3) | 1.1 (0.8, 1.5) | 1.0 (0.8, 1.3) | 1.0 (0.8, 1.4) | 0.0010 |
| TCH (mmol/L) | 1.9 (1.3, 2.6) | 1.8 (1.2, 2.5) | 1.9 (1.3, 2.6) | 2.0 (1.5, 2.5) | 1.9 (1.2, 3.0) | 0.13 |
| Creatinine (μmol/L) | 78.0 (59.0, 126.5) | 96.0 (62.0, 180.8) | 78.0 (59.0, 128.8) | 70.0 (56.0, 97.0) | 180.0 (79.0, 277.0) | <0.001 |
| Serum urea (mmol/L) | 6.7 (4.2, 11.8) | 8.3 (4.7, 15.5) | 6.4 (4.0, 11.7) | 5.7 (4.0, 9.2) | 13.0 (6.3, 22.2) | <0.001 |
| Sodium (mmol/L) | 136.3 ± 6.3 | 136.2 ± 6.6 | 136.1 ± 6.0 | 137.3 ± 5.2 | 138.6 ± 7.8 | <0.001 |
| C-reactive protein (mg/L) | 14.3 (7.6, 27.7) | 14.0 (7.7, 29.0) | 14.5 (8.5, 27.8) | 13.9 (8.5, 25.3) | 14.5 (8.1, 30.2) | 0.83 |
| White blood cell count<br>(10<sup>9</sup>/L) | 7.2 (4.8, 11.2) | 7.7 (5.1, 11.9) | 7.1 (4.9, 11.0) | 7.3 (4.8, 11.5) | 10.4 (6.7, 14.1) | <0.001 |
| Neutrophil count (10<sup>9</sup>/L) | 5.3 (3.2, 8.9) | 5.9 (3.6, 9.5) | 5.3 (3.3, 8.8) | 5.3 (3.2, 9.1) | 8.6 (5.0, 11.9) | <0.001 |
| Haemoglobin (g/L) | 100.9 ± 28.3 | 99.6 ± 27.6 | 103.3 ± 27.0 | 105.6 ± 24.9 | 91.2 ± 27.5 | <0.001 |
| Haematocrit (%) | 29.0 ± 8.0 | 28.8 ± 8.1 | 29.6 ± 7.7 | 30.4 ± 7.5 | 26.8 ± 8.0 | <0.001 |
| Platelet count (10<sup>9</sup>/L) | 71.0 (43.0, 118.0) | 67.0 (41.5, 111.5) | 74.0 (44.8, 119.3) | 70.0 (44.0, 110.0) | 55.0 (34.0, 92.0) | 0.0037 |
| INR | 2.2 (1.7, 2.8) | 2.5 (1.9, 3.1) | 2.1 (1.7, 2.7) | 2.1 (1.8, 2.7) | 2.4 (1.8, 3.2) | <0.001 |
| Scores |  |  |  |  |  |  |
| A-TANGO ACLF-CRP | 42.6 ± 6.7 | 44.5 ± 7.0 | 42.0 ± 7.0 | 42.3 ± 7.4 | 53.3 ± 6.9 | <0.001 |
| A-TANGO ACLF-WBC | 44.0 ± 6.8 | 46.0 ± 7.2 | 43.3 ± 7.2 | 44.2 ± 8.0 | 54.8 ± 6.8 | <0.001 |
| COSSH-ACLF | 6.9 ± 1.5 | 7.3 ± 1.7 | 6.8 ± 1.5 | 6.8 ± 1.5 | 6.8 ± 1.5 | <0.001 |
| COSSH-ACLF II | 7.7 ± 1.1 | 8.1 ± 1.1 | 7.7 ± 1.1 | 7.8 ± 1.2 | 8.7 ± 1.4 | <0.001 |
| CLIF-C ACLF | 45.6 ± 8.6 | 48.7 ± 8.3 | 45.6 ± 8.4 | 45.9 ± 9.6 | 58.5 ± 8.3 | <0.001 |
| A-TANGO OF | 10.0 (9.0, 11.0) | 10.0 (9.0, 12.0) | 10.0 (9.0, 11.0) | 10.0 (8.0, 12.0) | 13.0 (12.0, 15.0) | <0.001 |
| CLIF-OF | 10.0 (9.0, 11.0) | 10.0 (10.0, 11.0) | 10.0 (8.0, 11.0) | 9.0 (8.0, 11.0) | 13.0 (11.0, 14.0) | <0.001 |
| MELD-Na | 25.8 ± 7.8 | 28.5 ± 7.5 | 26.3 ± 7.1 | 25.5 ± 7.2 | 30.1 ± 10.0 | <0.001 |
| Transplant-free mortality (no.) |  |  |  |  |  |  |
| 28 days | 32.5% (305) | 41.5% (233) | 31.6% (309) | 32.4% (104) | 62.9% (39) | <0.001 |
| 90 days | 49.0% (423) | 57.8% (297) | 48.6% (439) | 43.8% (131) | 67.9% (40) | <0.001 |

Among 3,370 hospitalized patients with acutely decompensated cirrhosis, ACLF was diagnosed according to five different models. P values for comparisons among ACLF groups defined by the five diagnostic models. ACLF, acute-on-chronic liver failure; ALB, albumin; ALP, alkaline phosphatase; ALT, alanine aminotransferase; AST, aspartate aminotransferase; APASL, Asian Pacific Association for the Study of the Liver; CLIF-C, Chronic Liver Failure Consortium; COSSH, Chinese Group on the Study of Severe Hepatitis B; EASL-CLIF, European Association for the Study of the Liver-Chronic Liver Failure Consortium; HBV, hepatitis B virus; INR, international normalised ratio; MELD-Na, model for end-stage liver disease-sodium; NACSELD, North American Consortium for the Study of End-Stage Liver Disease; OF, organ failure; TB, total bilirubin; TCH, total cholesterol; TG, triglycerides.

Among patients classified as no ACLF by each diagnostic model, non-negligible short-term mortality was still observed. Specifically, 28–day LT-free mortality among no ACLF patients was 3.8% for A-TANGO, 6.2% for EASL-CLIF, 3.7% for COSSH-ACLF, 10.4% for APASL-ACLF, and 11.7% for NACSELD-ACLF, with corresponding 90–day LT-free mortality of 8.1%, 12.3%, 7.4%, 17.9%, and 19.7%. These findings indicate that a substantial proportion of patients with clinically meaningful short-term risk were not captured by stricter diagnostic models. Detailed clinical characteristics of no ACLF patients are provided in Supplemental Table S4.

### Clinical phenotypes of single-positive ACLF: A-TANGO vs COSSH-ACLF

Given the best discriminative diagnostic ability of the A-TANGO and COSSH-ACLF diagnostic models, a focused comparison was performed between mutually exclusive single-positive ACLF patients who met the A-TANGO definition but did not meet COSSH-ACLF (A-TANGO⁺/COSSH⁻) and those who met the COSSH-ACLF definition but did not meet A-TANGO (A-TANGO⁻/COSSH⁺) (Table 3). Compared with the A-TANGO⁻/COSSH⁺ group, the A-TANGO⁺/COSSH⁻ group more frequently presented with HE (21.4% vs 10.1%; P=0.0019) and gastrointestinal bleeding (26.2% vs 8.7%; P <0.001), but had lower rates of ascites (64.2% vs 73.6%; P=0.044) and infection (55.1% vs 66.8%; P=0.017). Etiologic distributions were similar between groups. In contrast, the A-TANGO⁻/COSSH⁺ group exhibited more pronounced hepatocellular injury and cholestasis, with higher alanine aminotransferase, aspartate aminotransferase, alkaline phosphatase, and bilirubin levels (all P <0.001), together with lower hemoglobin and hematocrit, higher platelet counts, and modestly higher CRP levels. Despite these phenotypic differences, short-term LT-free mortality was comparable between the A-TANGO⁺/COSSH⁻ and A-TANGO⁻/COSSH⁺ groups at 28 days (10.8% vs 10.7%; P=0.97) and 90 days (18.8% vs 23.9%; P=0.28), supporting an intermediate short-term risk profile among single-positive, model-exclusive patients.

**Table 3.** Characteristics of A-TANGO^+^ / COSSH^-^ and A-TANGO^-^ / COSSH^+^ groups.

| Characteristic | A-TANGO<sup>+</sup> / COSSH<sup>-</sup> (n=187) | A-TANGO<sup>-</sup> / COSSH<sup>+</sup> (n=208) | Total (n=395) | P value |
| --- | --- | --- | --- | --- |
| Age (years) | 55 ± 12 | 53 ± 11 | 54 ± 12 | 0.051 |
| Male (no.) | 70.6% (132) | 76.9% (160) | 73.9% (292) | 0.15 |
| Complication (no.) |  |  |  |  |
| Hepatic encephalopathy | 21.4% (40) | 10.1% (21) | 15.4% (61) | 0.0019 |
| Gastrointestinal bleeding | 26.2% (49) | 8.7% (18) | 17.0% (67) | <0.001 |
| Ascites | 64.2% (120) | 73.6% (153) | 69.1% (273) | 0.044 |
| Infection | 55.1% (103) | 66.8% (139) | 61.3% (242) | 0.017 |
| Etiologies (no.) |  |  |  |  |
| HBV related | 52.9% (99) | 52.9% (110) | 52.9% (209) | 0.99 |
| Alcohol related | 26.2% (49) | 32.7% (68) | 29.6% (117) | 0.16 |
| Autoimmune related | 9.6% (18) | 13.9% (29) | 11.9% (47) | 0.19 |
| Others | 17.6% (33) | 13.0% (27) | 15.2% (60) | 0.20 |
| Laboratory data |  |  |  |  |
| ALT (U/L) | 35.0 (21.0, 80.5) | 83.0 (41.8, 244.0) | 55.0 (26.0, 163.5) | <0.001 |
| AST (U/L) | 58.0 (33.0, 113.5) | 133.0 (75.0, 232.5) | 92.0 (46.0, 179.8) | <0.001 |
| ALP (U/L) | 106.5 (80.8, 152.9) | 145.0 (114.0, 187.0) | 128.0 (93.0, 173.0) | <0.001 |
| ALB (g/L) | 28.3 ± 4.6 | 28.9 ± 4.6 | 28.6 ± 4.6 | 0.17 |
| TB (μmol/L) | 135.7 (65.5, 193.7) | 266.2 (237.9, 294.1) | 231.8 (143.45, 289.8) | <0.001 |
| TG (mmol/L) | 0.9 (0.6, 1.4) | 1.2 (1.0, 1.6) | 1.1 (0.8, 1.6) | <0.001 |
| TCH (mmol/L) | 2.2 (1.7, 2.8) | 2.1 (1.7, 2.7) | 2.1 (1.7, 2.7) | 0.97 |
| Creatinine (μmol/L) | 68.0 (57.0, 85.0) | 69.0 (56.0, 87.0) | 69.0 (56.0, 85.1) | 0.95 |
| Serum urea (mmol/L) | 5.9 (4.1, 8.9) | 5.0 (3.6, 7.5) | 5.5 (3.8, 8.1) | 0.019 |
| Sodium (mmol/L) | 137.0 (135.0, 137.0) | 137.0 (133.1, 139.0) | 137.0 (134.0, 137.0) | 0.005 |
| C-reactive protein (mg/L) | 10.7 (5.0, 24.2) | 14.3 (9.6, 24.7) | 13.6 (7.0, 24.7) | 0.008 |
| White blood cell count (10<sup>9</sup>/L) | 5.7 (3.3, 9.1) | 6.2 (4.5, 8.5) | 5.9 (4.0, 8.7) | 0.099 |
| Neutrophil count (10<sup>9</sup>/L) | 3.8 (2.0, 6.8) | 4.1 (3.0, 6.1) | 4.0 (2.6, 6.5) | 0.11 |
| Haemoglobin (g/L) | 92.7 ± 30.1 | 106.8 ± 23.6 | 100.1 ± 27.8 | <0.001 |
| Haematocrit (%) | 27.2 ± 8.1 | 30.7 ± 6.7 | 29.0 ± 7.6 | <0.001 |
| Platelet count (10<sup>9</sup>/L) | 57.0 (39.5, 113.5) | 84.5 (53.0, 122.3) | 73.0 (44.5, 119.0) | 0.0020 |
| INR | 1.7 (1.4, 2.4) | 1.8 (1.6, 2.0) | 1.8 (1.5, 2.1) | 0.34 |
| Scores |  |  |  |  |
| A-TANGO ACLF-CRP | 39.4 ± 5.1 | 36.4 ± 4.4 | 37.8 ± 5.0 | <0.001 |
| A-TANGO ACLF-WBC | 40.5 ± 4.8 | 37.0 ± 4.3 | 38.7 ± 4.9 | <0.001 |
| COSSH-ACLF | 6.0 ± 0.6 | 5.7 ± 0.4 | 5.8 ± 0.6 | <0.001 |
| COSSH-ACLF II | 6.9 ± 1.0 | 6.9 ± 0.6 | 6.9 ± 0.8 | 0.75 |
| CLIF-C ACLF | 40.4 ± 7.0 | 40.5 ± 5.6 | 40.4 ± 6.3 | 0.98 |
| A-TANGO OF | 9.0 (8.0, 9.0) | 8.0 (8.0, 8.0) | 8.0 (8.0, 9.0) | <0.001 |
| CLIF-OF | 8.0 (8.0, 9.0) | 8.0 (8.0, 9.0) | 8.0 (8.0, 9.0) | 0.42 |
| MELD-Na | 18.3 ± 6.8 | 21.6 ± 4.4 | 20.0 ± 5.9 | <0.001 |
| Transplant-free mortality (no.) |  |  |  |  |
| 28 days | 10.8% (17) | 10.7% (21) | 10.8% (38) | 0.97 |
| 90 days | 18.8% (26) | 23.9% (43) | 21.7% (69) | 0.28 |

P values for comparisons between A-TANGO<sup>+</sup> / COSSH<sup>-</sup> and A-TANGO<sup>-</sup> / COSSH<sup>+</sup> groups. ACLF, acute-on-chronic liver failure; ALB, albumin; ALP, alkaline phosphatase; ALT, alanine aminotransferase; AST, aspartate aminotransferase; APASL, Asian Pacific Association for the Study of the Liver; CLIF-C, Chronic Liver Failure Consortium; COSSH, Chinese Group on the Study of Severe Hepatitis B; EASL-CLIF, European Association for the Study of the Liver-Chronic Liver Failure Consortium; HBV, hepatitis B virus; INR, international normalised ratio; MELD-Na, model for end-stage liver disease-sodium; NACSELD, North American Consortium for the Study of End-Stage Liver Disease; OF, organ failure; TB, total bilirubin; TCH, total cholesterol; TG, triglycerides.

### Discriminative performance of prognostic scores for 28–day LT-free mortality

In the overall cohort, the COSSH-ACLF II score demonstrated the highest discrimination for 28-day mortality (C-index 0.857, 95% CI 0.836–0.877), with performance comparable to the COSSH-ACLF score (0.848, 95% CI 0.827–0.870). Both scores outperformed the A-TANGO OF, CLIF-OF, and MELD-Na scores, whereas the NACSELD-ACLF score showed markedly lower discrimination (0.547, 95% CI 0.530–0.564) (Table 4). Among the 388 patients with available AARC data, most prognostic scores demonstrated broadly similar discrimination. COSSH-ACLF II, COSSH-ACLF, and A-TANGO OF scores yielded comparable C-index estimates, while the NACSELD-ACLF score again showed substantially poorer performance (Supplemental Table S5). Discriminative performance differed across model-defined ACLF populations (Supplemental Tables S6–S10). COSSH-based scores (COSSH-ACLF II and COSSH-ACLF) were often among the highest-performing scores, whereas A-TANGO-based scores (A-TANGO OF score, A-TANGO ACLF-WBC and A-TANGO ACLF-CRP) showed discrimination comparable to CLIF-based scores (CLIF-OF score and CLIF-C ACLF score) in most analyses. NACSELD-ACLF consistently demonstrated the lowest discrimination across strata.

**Table 4.** Pairwise comparison of discrimination performance (concordance index and integrated discrimination improvement) of prognostic scores for predicting 28–day liver transplant–free mortality in the entire cohort.

| C-index of various prognostic scores |  |  |  |  |  |  |
| --- | --- | --- | --- | --- | --- | --- |
| Score | CLIF-OF | A-TANGO OF | COSSH-ACLF | COSSH-ACLF II | NACSELD-ACLF | MELD-Na |
| CLIF-OF | 0.827 (0.804–0.851) | – | – | – | – | – |
| A-TANGO OF | 0.41 | 0.832 (0.810–0.854) | – | – | – | – |
| COSSH-ACLF | <0.001 | 0.0017 | 0.848 (0.827–0.870) | – | – | – |
| COSSH-ACLF II | 0.0011 | 0.0041 | 0.23 | 0.857 (0.836–0.877) | – | – |
| NACSELD-ACLF | <0.001 | <0.001 | <0.001 | <0.001 | 0.547 (0.530–0.564) | – |
| MELD-Na | 0.65 | 0.32 | 0.0034 | <0.001 | <0.001 | 0.823 (0.801–0.845) |
| Diagonal: C-index with 95% CI; off-diagonal: P values for pairwise comparison |  |  |  |  |  |  |
| IDI of various prognostic scores |  |  |  |  |  |  |
| Score | CLIF-OF | A-TANGO OF | COSSH-ACLF | COSSH-ACLF II | NACSELD-ACLF | MELD-Na |
| CLIF-OF | Reference | – | – | – | – | – |
| A-TANGO OF | 0.81% (-1.21% to 2.93%) | Reference | – | – | – | – |
| COSSH-ACLF | 0.62% (-4.29% to 4.61%) | –0.19% (-5.20% to 4.06%) | Reference | – | – | – |
| COSSH-ACLF II | 4.09% (0.79% to 7.48%) ** | 3.28% (-0.02% to 6.46%) * | 3.47% (-0.31% to 7.67%) | Reference | – | – |
| NACSELD-ACLF | -21.03% (-25.50% to -17.00%)<br>*** | -21.84% (-26.13% to -17.70%)<br>*** | -21.65% (-27.79% to -15.56%)<br>*** | -25.12% (-29.73% to -20.37%)<br>*** | Reference | — |
| MELD-Na | -3.73% (-7.21% to -0.26%) ** | -4.54% (-8.13% to -1.26%) ** | -4.35% (-8.98% to 0.36%) * | -7.83% (-10.81% to -4.91%)<br>*** | 17.30% (12.99% to 21.51%)<br>*** | Reference |
| Off-diagonal: IDI and 95% CI compared with reference score in each column; $P \leq 0.05$ is indicated by *, $P \leq 0.01$ by **, and $P \leq 0.001$ by *** | | | | | | |

ACLF, acute-on-chronic liver failure; C-index, concordance index; CLIF-OF, Chronic Liver Failure-organ failure; COSSH-ACLF, Chinese Group on the Study of Severe Hepatitis B-ACLF; COSSH-ACLF II, Chinese Group on the Study of Severe Hepatitis B-ACLF II; IDI, integrated discrimination improvement; MELD-Na, Model for End-Stage Liver Disease-sodium; NACSELD-ACLF, North American Consortium for the Study of End-Stage Liver Disease-ACLF.

Pairwise IDI analyses further supported these findings. Using CLIF-OF score as the reference, the COSSH-ACLF II score provided significant incremental discrimination (IDI 4.09%), whereas COSSH-ACLF and A-TANGO OF scores showed no meaningful improvement. MELD-Na and NACSELD-ACLF scores were associated with reduced discrimination, with the largest deterioration observed for NACSELD-ACLF score (Table 4). In the AARC subset, COSSH-ACLF II score showed the largest numerical IDI versus CLIF-OF score, while NACSELD-ACLF score showed a significant deterioration (Supplemental Table S11). Across model-specific ACLF populations, the COSSH-ACLF II score frequently ranked among the top-performing models, whereas the NACSELD-ACLF score showed predominantly negative IDI values, suggesting poorer reclassification performance (Supplemental Tables S12–S16).

### Calibration and decision-curve analysis of prognostic scores for 28–day LT-free mortality

Calibration analyses demonstrated overall agreement between predicted and observed 28-day mortality across prognostic scores, with low mean absolute errors (Figure 2A). COSSH-ACLF II score showed near-ideal calibration across the risk spectrum, whereas A-TANGO OF and CLIF-OF scores also demonstrated close alignment between predicted and observed risk. COSSH-ACLF score exhibited generally acceptable calibration but showed modest underestimation at higher predicted probabilities. Similarly, MELD-Na score tended to underestimate mortality in higher-risk ranges despite low average error. In contrast, meaningful calibration assessment for NACSELD-ACLF score was limited by its binary risk structure.

**Figure 2.**
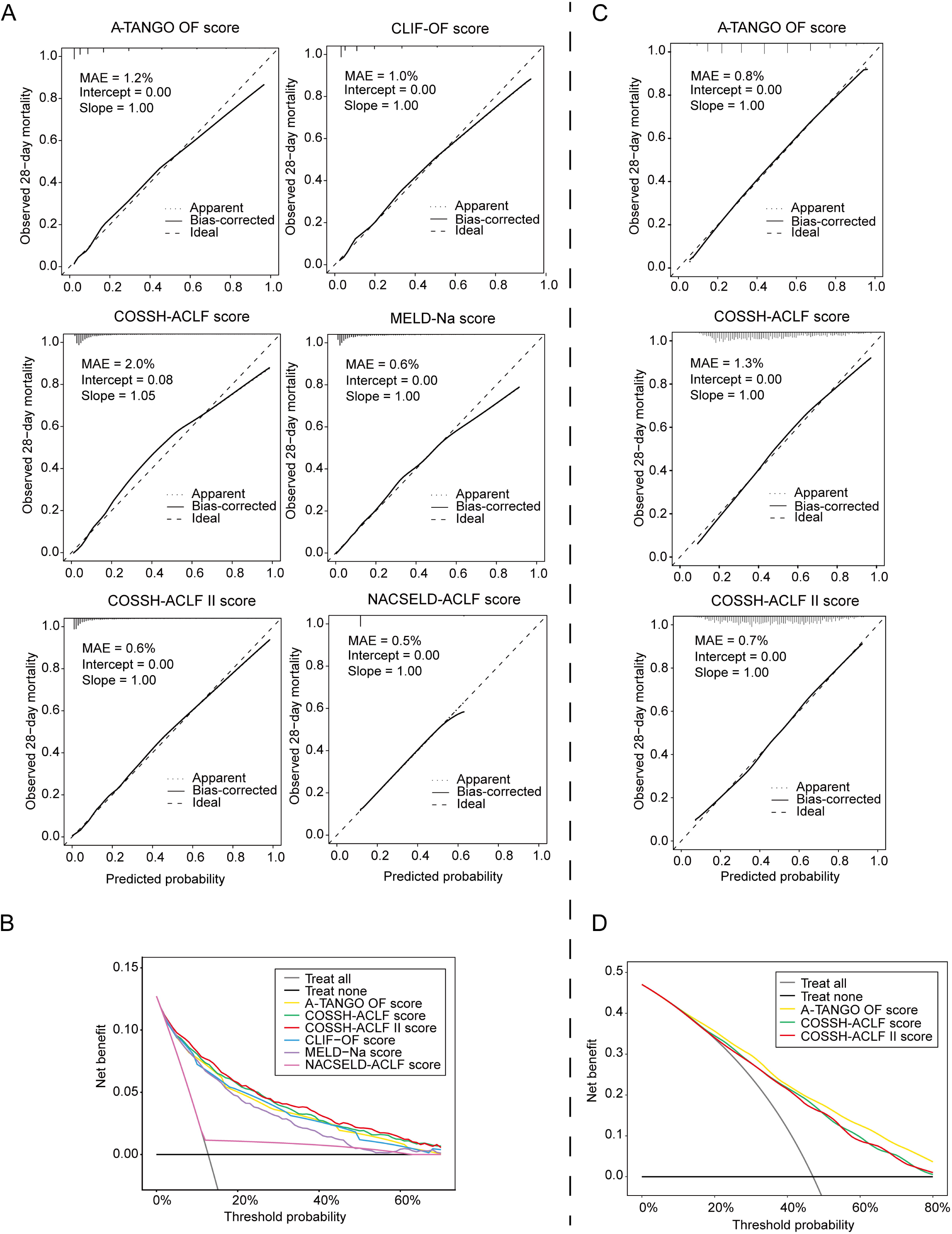
Prognostic performance of prognostic scores for 28-day liver transplant-free mortality. (A) Calibration in the COSSH cohort (n=3,370). Predicted risk is plotted against observed mortality; the dashed line indicates perfect calibration. Internal validation used bootstrap resampling (1,000 iterations). (B) Decision curve analysis in the COSSH cohort. (C) External calibration in the Indian Ambi-Spective cohort. (D) Decision curve analysis in the Indian Ambi-Spective cohort. ACLF, acute-on-chronic liver failure; CLIF, Chronic Liver Failure; COSSH, Chinese Group on the Study of Severe Hepatitis B; MELD-Na, model for end-stage liver disease-sodium; NACSELD, North American Consortium for the Study of End-Stage Liver Disease; OF, organ failure.

Decision-curve analysis demonstrated differences in clinical utility among prognostic scores (Figure 2B). The COSSH-ACLF II score yielded the highest net benefit across a broad range of threshold probabilities, with COSSH-ACLF and A-TANGO OF scores showing closely comparable performance. CLIF-OF and MELD-Na scores provided modest net benefit but remained superior to the treat-all and treat-none strategies. In contrast, NACSELD-ACLF score showed substantially lower net benefit across thresholds.

### Comparison of COSSH-ACLF and A-TANGO Diagnostic Models in an External Indian Cohort

As the COSSH-ACLF diagnostic model was developed in the COSSH cohort, we assessed the performance of the COSSH-ACLF and A-TANGO diagnostic models and their corresponding prognostic scores in an external, independent study population, the Ambi-Spective cohort from India (n=2,055). Despite differences in overall cohort characteristics compared with the COSSH cohort, patients classified as ACLF by the COSSH-ACLF (n=1,697) and A-TANGO (n=1,725) models exhibited highly comparable baseline profiles (Supplemental Table S17). However, no meaningful differences were observed in demographics, precipitating events, etiologies, complications, laboratory findings, or disease severity scores between A-TANGO and COSSH defined ACLF. Consistent with these similarities, 28- and 90-day LT-free mortality were nearly identical between groups, indicating substantial overlap in the high-risk populations identified by the two diagnostic models. In ROC space, the COSSH-ACLF and A-TANGO models clustered closely, indicating similar discriminative ability for 28-day LT-free mortality (Figure 1D). Both models demonstrated high sensitivity and strong negative predictive values, suggesting utility for early risk identification and reliable exclusion of short-term mortality (Figure 1E). Kaplan–Meier analyses showed graded survival across ACLF severity for both models, with progressively lower survival observed with increasing ACLF grade (Supplemental Figure S1B). The A-TANGO grading was confirmed to achieve refined risk stratification relative to COSSH-grades for 28-day LT-free mortality (NRI=17.1%), primarily through upward reclassification of non-survivors (Supplemental Table S18).

In this external cohort, the prognostic scores demonstrated similar discriminative performance. A-TANGO OF score achieved the highest C-index, followed by COSSH-ACLF II score, with both modestly exceeding COSSH-ACLF score. Pairwise comparisons showed no significant difference between A-TANGO OF and COSSH-ACLF II scores (Supplemental Table S19), whereas IDI analyses supported incremental discrimination with A-TANGO OF score (Supplemental Table S20). All models were well calibrated; COSSH-ACLF II score showed the closest agreement between predicted and observed risk, whereas A-TANGO OF score yielded the greatest net benefit on decision-curve analysis (Figures 2C, D). These findings suggest consistent performance of COSSH-ACLF and A-TANGO across populations, with COSSH-ACLF II score demonstrating slightly stronger statistical calibration and A-TANGO OF score offering greater potential clinical utility, thereby supporting complementary roles in risk assessment.

### Risk stratification through integrated application of the A-TANGO and COSSH-ACLF diagnostic models

Given the substantial overlap but imperfect concordance between the A-TANGO and COSSH-ACLF diagnostic models, we evaluated whether their combined application could enable more refined risk stratification in the COSSH cohort. Concurrent use of the two diagnostic models stratified patients according to concordant and discordant classifications. Patients positive by both models (A-TANGO^+^/COSSH^+^) represented the highest-risk subgroup, whereas those positive by only one model experienced clinically meaningful 28-day LT-free mortality (∼11%). In contrast, patients negative by both models showed very low mortality risk (3.1%) (Figure 3A). This integrated approach stratified the cohort into three clinically meaningful risk groups—high, intermediate, and low risk. Kaplan–Meier analyses demonstrated clear separation of survival across strata, with significant differences observed in both overall and pairwise comparisons (log-rank P <0.0001 for all) (Figure 3B), demonstrating the clinical relevance of this integrated model. Consistent with these observations, NRI analyses showed modest but directionally favorable reclassification for double-positive patients compared with either model alone (NRI: 0.9% vs A-TANGO; 1.2% vs COSSH-ACLF) (Supplemental Table S21). Figure 3C illustrates a pragmatic two-step strategy in which concurrent model application first identifies concordant and discordant patterns, which are subsequently consolidated into three risk categories to guide escalation of clinical management. Among patients in the high-risk subgroup (A-TANGO^+^/COSSH^+^, n=947), the COSSH-ACLF II score showed strong performance across discrimination, incremental discrimination, reclassification, calibration, and clinical utility, supporting its role in refining prognostic assessment within this population (Supplemental Tables S22 and S23; Supplemental Figures S2 and S3).

**Figure 3.**
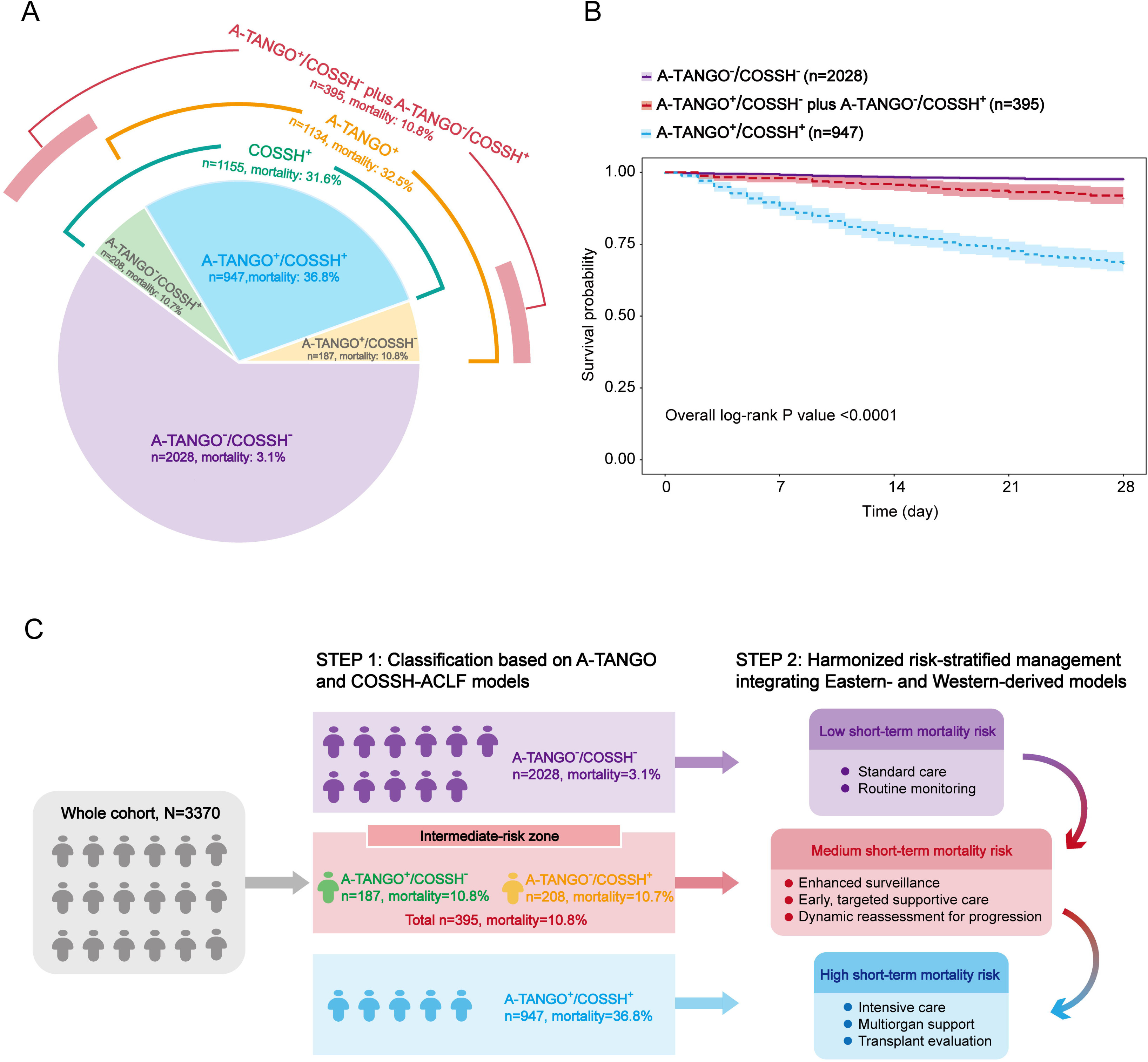
Risk stratification based on A-TANGO and COSSH-ACLF models in the COSSH cohort. (A) Overlap between the two models. (B) Kaplan–Meier curves for 28-day survival across three risk groups (log-rank P<0.0001 for all comparisons). (C) Stepwise risk stratification framework based on their combined application. ACLF, acute-on-chronic liver failure; APASL, Asian Pacific Association for the Study of the Liver; COSSH, Chinese Group on the Study of Severe Hepatitis B; EASL-CLIF, European Association for the Study of the Liver-Chronic Liver Failure Consortium; LT, liver transplantation; NACSELD, North American Consortium for the Study of End-Stage Liver Disease; NPV, negative predictive value; PPV, positive predictive value.

Given the distinct clinical profile observed in patients classified as medium risk by discordant A-TANGO and COSSH-ACLF diagnostic models, we further explored determinants of outcome within this subgroup (Table 5). Univariable and multivariable Cox proportional hazards regression analyses were performed to identify factors associated with 90-day LT-free mortality. Variables significant in univariable analyses, together with clinically relevant covariates, were entered into the multivariable model. In this intermediate-risk group, most conventional severity-related variables, including markers of organ dysfunction commonly incorporated into ACLF prognostic scores, were not independently associated with outcome. Instead, multivariable analysis identified total cholesterol and white blood cell count as independent predictors of mortality. Sensitivity analyses substituting neutrophil count for white blood cell count yielded similar results, reinforcing the stability of these associations (Supplemental Table S24). Together, these findings suggest that, in contrast to high- and low-risk groups, the intermediate-risk subgroup represents a distinct prognostic profile, with outcomes appearing more closely associated with inflammatory and metabolic markers than with traditional indicators of organ dysfunction.

**Table 5.** Univariable and multivariable Cox analysis for 90–day liver transplant-free mortality in the intermediate-risk group (n=395).

| Parameter | Univariable HR (95% CI) | P value | Multivariable HR (95% CI) | P value |
| --- | --- | --- | --- | --- |
| Male (yes vs no) | 0.54 (0.31–0.92) | 0.025 | 0.56 (0.31–1.01) | 0.053 |
| Age (years) | 1.02 (1.00–1.05) | 0.058 | 1.02 (0.99–1.05) | 0.12 |
| Hepatic encephalopathy (yes vs no) | 1.10 (0.52–2.32) | 0.81 | - | - |
| Gastrointestinal bleeding (yes vs no) | 1.54 (0.83–2.87) | 0.17 | - | - |
| Ascites (yes vs no) | 0.81 (0.47–1.40) | 0.45 | - | - |
| Infection (yes vs no) | 0.98 (0.57–1.68) | 0.93 | - | - |
| HBV related (yes vs no) | 0.68 (0.40–1.15) | 0.15 | - | - |
| Alcohol related (yes vs no) | 0.65 (0.34–1.23) | 0.18 | - | - |
| Autoimmune related (yes vs no) | 1.91 (0.96–3.79) | 0.064 | - | - |
| ALT (U/L) | 1.00 (1.00–1.00) | 0.66 | - | - |
| AST (U/L) | 1.00 (1.00–1.00) | 0.054 | - | - |
| ALP (U/L) | 1.00 (0.99–1.00) | 0.15 | - | - |
| ALB (g/L) | 0.99 (0.94–1.05) | 0.81 | - | - |
| TB (μmol/L) | 1.00 (1.00–1.00) | 0.14 | - | - |
| TG (mmol/L) | 0.78 (0.51–1.19) | 0.25 | - | - |
| TCH (mmol/L) | 0.61 (0.43–0.88) | 0.0077 | 0.62 (0.43–0.88) | 0.0085 |
| Creatinine (μmol/L) | 1.00 (0.99–1.01) | 0.60 | - | - |
| Serum urea (mmol/L) | 1.02 (0.97–1.08) | 0.38 | - | - |
| Sodium (mmol/L) | 0.94 (0.90–0.99) | 0.012 | 0.96 (0.91–1.01) | 0.12 |
| C-reactive protein (mg/L) | 1.00 (0.98–1.00) | 0.48 | - | - |
| White blood cell count (10<sup>9</sup>/L) | 1.07 (1.01–1.12) | 0.017 | 1.07 (1.01–1.15) | 0.032 |
| Neutrophil count (10<sup>9</sup>/L) | 1.08 (1.02–1.14) | 0.0095 | - | - |
| Haemoglobin (g/L) | 1.00 (0.99–1.01) | 0.95 | - | - |
| Haematocrit (%) | 1.00 (0.96–1.03) | 0.89 | - | - |
| Platelet count (10<sup>9</sup>/L) | 1.00 (0.99–1.00) | 0.050 | - | - |
| INR | 1.44 (0.96–2.16) | 0.082 | - | - |

ALB, albumin; ALP, alkaline phosphatase; ALT, alanine aminotransferase; AST, aspartate aminotransferase; HBV, hepatitis B virus; INR, international normalised ratio; TB, total bilirubin; TCH, total cholesterol; TG, triglycerides.

## Discussion

This large multicenter, multinational study provides important insights into the evolving landscape of ACLF diagnosis and risk assessment, informing approaches to harmonization. Despite substantial conceptual differences, geographical origins, patient populations and standard of clinical care among contemporary diagnostic models, our findings suggest a notable degree of alignment between outcome-driven criteria, particularly A-TANGO and COSSH-ACLF diagnostic models, in identifying patients with ACLF. At the same time, discordant classification between these two models revealed a clinically meaningful intermediate-risk population, reinforcing the notion that AD represents a continuum of disease severity rather than a binary ACLF versus non-ACLF presentation, and supporting a data-driven framework for harmonized risk stratification.

Among the diagnostic models, the A-TANGO diagnostic model demonstrated refined risk stratification and a strong ability to identify high-risk patients. Developed using prospective cohorts of 3,896 patients from Europe^1,25^ and Latin America^26^ and validated in large cohorts from India and China, it incorporates recalibrated organ failure sub scores, combines HE grades 0 and 1, and adds an additional ACLF grade. These refinements enable a more stepwise increase in 28–day mortality across ACLF grades, reduces the heterogeneity seen in EASL-CLIF grade 3, and improves discrimination of patients at extreme risk. The EASL-CLIF diagnostic model was the first to define ACLF based on organ failure assessment and has since been extensively validated.^1^ However, several inherent limitations such as the cut-offs used to identify organ failures, diagnosis of early HE and renal failure have been shown to reduce its sensitivity in identifying ACLF.^15^ In addition, the broad mortality range within EASL-CLIF ACLF grade 3 increases patient heterogeneity, thereby limiting its ability to discriminate individuals with extremely high mortality.

The COSSH-ACLF diagnostic model also identified a large proportion of high-risk patients with good short-term mortality discrimination. Its diagnostic model incorporates patients with total bilirubin ≥12 mg/dL and an INR ≥1.5 as ACLF grade 1, thereby capturing individuals with significant hepatic dysfunction at an early stage.^9^ This broader inclusion criterion enhances diagnostic sensitivity and results in performance comparable to the A-TANGO diagnostic model in identifying patients at risk of poor outcomes. Although its grading structure effectively reflects the spectrum of organ dysfunction, it provides slightly less granularity than the A-TANGO diagnostic model in distinguishing the highest-risk subgroups. The APASL-ACLF diagnostic model emphasizes hepatic and coagulation failure as core diagnostic components, requiring jaundice and coagulopathy following an acute hepatic insult.^13,27^ However, its focus on excluding patients with prior decompensation is of limited prognostic value, as short-term mortality remains comparable between those with and without such a history.^28^ Moreover, its limited consideration of extrahepatic organ failures further contributes to the model missing a broader spectrum of high-risk patients and ultimately reduces its prognostic precision. The NACSELD-ACLF diagnostic model focuses exclusively on extrahepatic organ failures, defining ACLF only when two or more non-hepatic organ failures are present.^12,29^ This stringent threshold identifies a small subset of critically ill patients with very high short-term mortality but fails to detect those with isolated hepatic failure or single-organ dysfunction who are still at considerable risk, limiting its diagnostic sensitivity and prognostic discrimination.

Within the overall COSSH cohort, A-TANGO and COSSH-ACLF diagnostic models identified the largest proportion of patients at substantial short-term risk of mortality, supporting the value of outcome-anchored approaches to ACLF definition. Importantly, this pattern was reproduced in the external cohort, where both models demonstrated comparable diagnostic performance despite differences in underlying patient populations. Consistency across geographically distinct settings suggests that data-driven frameworks may more reliably capture clinically meaningful risk than definitions derived primarily through consensus.

In addition, our findings indicate that the A-TANGO and COSSH-ACLF diagnostic models should not be viewed as competing frameworks but rather as complementary tools that capture different dimensions of short-term risk, which may indicate first steps towards harmonization. Application of either A-TANGO or COSSH-ACLF alone identified overlapping yet non-identical subsets of patients, whereas their parallel application enabled a more refined risk stratification by delineating a medium-risk group that was not consistently classified by either model alone. On this basis, our results support a stepwise, risk-oriented diagnostic approach in which concordant positivity across these two models identifies patients at the highest short-term risk, while discordant or isolated positivity corresponds to a medium-risk state. Such a framework may facilitate more rational resource allocation by prioritizing intensive management for high-risk patients while avoiding unnecessary escalation in lower-risk individuals. It is also important to consider the etiological context in which these models were developed. The COSSH-ACLF diagnostic model was derived mainly from hepatitis B–related populations, whereas A-TANGO was developed in predominantly no hepatitis B–related cohorts. In settings where a specific etiology predominates, this distinction may inform model selection. However, the consistent performance of both models across diverse populations in our study suggests that they are broadly applicable, and their combined use may provide a pragmatic approach to capturing the full spectrum of short-term risk.

The prognostic analyses highlight the consistently strong performance of scores derived from the COSSH-ACLF and A-TANGO diagnostic models. While the COSSH-ACLF II score showed the highest discrimination within the COSSH cohort and the A-TANGO OF score performed better in the Ambi-Spective cohort, both model families demonstrated robust predictive ability across populations. These findings suggest that outcome-anchored model development can provide a durable foundation for short-term risk prediction despite geographic and clinical heterogeneity. Such a pattern is not unique to ACLF. In end-stage liver disease, successive refinements from MELD score^30^ to MELD-Na score^16^ and MELD 3.0 score^31^ similarly resulted in incremental gains in predictive accuracy, illustrating how structured recalibration can progressively enhance prognostic performance.

Several limitations warrant consideration. First, although the COSSH-ACLF framework was developed within the primary cohort, external validation mitigates concerns of derivation bias. Second, reliance on NRI and IDI metrics, while informative, must be interpreted cautiously given their dependence on risk categorization thresholds.^19,20^ Third, variability in hepatic encephalopathy grading and organ failure assessment may influence reproducibility across settings. Nonetheless, consistent discrimination, calibration, and decision-curve findings across cohorts support the robustness of our conclusions.

In summary, systematic comparison of five ACLF diagnostic models identified A-TANGO and COSSH-ACLF as particularly effective for short-term mortality risk stratification, while discordant classification between the two models delineated a clinically meaningful intermediate-risk population. Prognostic scores derived from these outcome-based diagnostic models demonstrated strong and reproducible predictive performance, with COSSH-ACLF II score achieving the highest discrimination in the COSSH cohort and A-TANGO OF score performing better in the external cohort. The concordance of diagnostic behavior together with the stability of prognostic performance across populations provides important context for interpreting ACLF risk. Together, these findings provide an evidence-based foundation for advancing discussions regarding global harmonization of ACLF definition while preserving the strengths of existing validated models.

## List of Abbreviations

AARC, APASL ACLF Research Consortium; ACLF, acute-on-chronic liver failure; ALB, albumin; ALP, alkaline phosphatase; ALT, alanine aminotransferase; APASL, Asian Pacific Association for the Study of the Liver; AST, aspartate aminotransferase; C-index, concordance index; CLIF-C, Chronic Liver Failure Consortium; COSSH, Chinese Group on the Study of Severe Hepatitis B; CRP, C-reactive protein; DCA, decision curve analysis; EASL-CLIF, European Association for the Study of the Liver–Chronic Liver Failure Consortium; HBV, hepatitis B virus; HE, hepatic encephalopathy; IDI, integrated discrimination improvement; INR, international normalised ratio; LT, liver transplantation; MELD-Na, model for end-stage liver disease–sodium; NACSELD, North American Consortium for the Study of End-Stage Liver Disease; NPV, negative predictive value; NRI, net reclassification improvement; OF, organ failure; PPV, positive predictive value; TB, total bilirubin; TCH, total cholesterol; TG, triglycerides; WBC, white blood cell count.

## Data Availability

All data are available from the corresponding author upon reasonable request.

## Acknowledgements

The authors thank all the physicians, nurses, and research staff involved in this study for their contributions to patient care and data collection.

