## Supplementary Materials for "A Comparison of Diagnostic Models and Prognostic Scores of ACLF: Towards Global Harmonization"

**Supplemental Table 1.** Diagnostic models of EASL-CLIF, COSSH-ACLF, A-TANGO, APASL-ACLF, and NACSELD-ACLF.

**Supplemental Table 2.** Calculation formula of prognostic scores.

**Supplemental Table 3.** Comprehensive comparison of five ACLF diagnostic models based on classification agreement.

**Supplemental Table 4.** Characteristics of no ACLF patients according to five diagnostic models.

**Supplemental Table 5.** Comparison of concordance index among various prognostic scores for predicting 28–day liver transplant-free mortality among patients with AARC data.

**Supplemental Table 6.** Comparison of concordance index among various prognostic scores for predicting 28–day liver transplant-free mortality according to the A-TANGO diagnostic model.

**Supplemental Table 7.** Comparison of concordance index among various prognostic scores for predicting 28–day liver transplant-free mortality according to the EASL-CLIF diagnostic model.

**Supplemental Table 8.** Comparison of concordance index among various prognostic scores for predicting 28–day liver transplant-free mortality according to the COSSH-ACLF diagnostic model.

**Supplemental Table 9.** Comparison of concordance index among various prognostic scores for predicting 28–day liver transplant-free mortality according to the APASL-ACLF diagnostic model.

**Supplemental Table 10.** Comparison of concordance index among various prognostic scores for predicting 28–day liver transplant-free mortality according to the NACSELD-ACLF diagnostic model.

**Supplemental Table 11.** Integrated discrimination improvement of prognostic scores for 28–day liver transplant-free mortality among patients with available AARC scores.

**Supplemental Table 12.** Integrated discrimination improvement of various prognostic scores for 28–day liver transplant-free mortality prediction according to the A-TANGO model.

**Supplemental Table 13.** Integrated discrimination improvement of various prognostic scores for 28–day liver transplant-free mortality prediction according to the EASL-CLIF model.

**Supplemental Table 14.** Integrated discrimination improvement of various prognostic scores for 28–day liver transplant-free mortality prediction according to the COSSH-ACLF model.

**Supplemental Table 15.** Integrated discrimination improvement of various prognostic scores for 28–day liver transplant-free mortality prediction according to the APASL-ACLF model.

**Supplemental Table 16.** Integrated discrimination improvement of various prognostic scores for 28–day liver transplant-free mortality prediction according to the NACSELD-ACLF model.

**Supplemental Table 17.** Clinical characteristics of patients with ACLF defined by the A-TANGO and COSSH-ACLF diagnostic models in the Ambi-Spective cohort from India.

**Supplemental Table 18.** Net reclassification improvement of A-TANGO versus COSSH-ACLF for predicting 28–day liver transplant-free mortality in the Ambi-Spective cohort from India.

**Supplemental Table 19.** Concordance index among various prognostic scores for predicting 28–day liver transplant-free mortality in the Ambi-Spective cohort from India.

**Supplemental Table 20.** Integrated discrimination improvement of various prognostic scores for 28–day liver transplant-free mortality prediction in the Ambi-Spective cohort from India.

**Supplemental Table 21.** Net reclassification improvement of the A-TANGO^+^/COSSH^+^ diagnostic model compared with the A-TANGO diagnostic model or COSSH diagnostic model only for 28–day liver transplant-free mortality.

**Supplemental Table 22.** Concordance index of various prognostic scores for 28–day liver transplant-free mortality prediction in the ATATGO^+^ /COSSH^+^ group.

**Supplemental Table 23.** Integrated discrimination improvement of various prognostic scores for 28–day liver transplant-free mortality prediction in the ATATGO^+^ /COSSH^+^ group.

**Supplemental Table 24.** Sensitivity analyses of multivariable Cox models for 90–day liver transplant-free mortality in the intermediate-risk group.

**Supplemental Figure 1.** Survival stratification according to ACLF grades defined by diferent diagnostic models.

**Supplemental Figure 2.** Calibration of prognostic scores for 28–day liver transplant-free mortality in the ATANGO^+^/COSSH^+^ group.

**Supplemental Figure 3.** Decision curve analysis of prognostic scores for 28–day liver transplant-free mortality in the ATANGO^+^/COSSH^+^ group.

**Supplemental Table 1.** Diagnostic models of EASL-CLIF, COSSH-ACLF, A-TANGO, APASL-ACLF, and NACSELD-ACLF.

| Characteristics | European Association for the Study of the Liver-Chronic Liver Failure Consortium (EASL-CLIF) | Chinese Group on the Study of Severe Hepatitis B (COSSH) | A-TANGO | Asian Pacific Association for the Study of the Liver (APASL) | North American Consortium for the Study of End-Stage Liver Disease (NACSELD) |
| --- | --- | --- | --- | --- | --- |
| Definition | The EASL-CLIF defines ACLF as a complicated syndrome that can develop in patients with acutely decompensated cirrhosis and is characterized by hepatic and/or extrahepatic organ failure and a high short-term mortality rate (≥ 15% at 28 days). | The COSSH-ACLF defines ACLF as a complicated syndrome associated with a high short-term mortality rate (≥ 15% at 28 days) that develops in patients with chronic liver disease, regardless of the presence of cirrhosis, and it is characterized by acute deterioration of liver function and hepatic and/or extrahepatic organ failure. | The A-TANGO defines ACLF as a complicated syndrome that can develop in patients with acutely decompensated cirrhosis and is characterized by hepatic and/or extrahepatic organ failure and a high short-term mortality rate (≥ 15% at 28 days). | The APASL-ACLF defined ACLF as a syndrome of acute liver function damage on the basis of known or unknown chronic liver disease. The definition mainly includes patients with compensated liver disease. | The NACSELD-ACLF defined ACLF as a severe clinical syndrome occurring in hospitalized patients with cirrhosis, characterized by the presence of multiple extrahepatic organ failures and an associated high short-term mortality risk. |
| Organ failure | Liver: Total bilirubin ≥ 12 mg/dL;  Kidney: Creatinine ≥ 2 mg/dL or use of RRT;  Coagulation: INR ≥ 2.5;  Brain: West Haven grade 3–4 HE or use of mechanical ventilation due to HE;  Circulation: Use of vasopressors;  Respiration: PaO2/FiO2 ≤ 200 or SpO2/FiO2 ≤ 214, or use of mechanical ventilation not due to HE. | Liver: Total bilirubin ≥ 12 mg/dL;  Kidney: Creatinine ≥ 2 mg/dL or use of RRT;  Coagulation: INR ≥ 2.5;  Brain: West Haven grade 3–4 HE or use of mechanical ventilation due to HE;  Circulation: Use of vasopressors;  Respiration: PaO2/FiO2 ≤ 200 or SpO2/FiO2 ≤ 214, or use of mechanical ventilation not due to HE. | Liver: Total bilirubin ≥ 20 mg/dL;  Kidney: Creatinine ≥ 2 mg/dL or use of RRT or AKI 1b, 2 or 3;  Coagulation: INR ≥ 2.2;  Brain: West Haven grade 3–4 HE or mechanical ventilation for the indication of cerebral failure;  Circulation: Use of vasopressors;  Respiration: PaO2/FiO2 ≤ 225 or SpO2/FiO2 ≤ 325, or mechanical ventilation for the indication of respiratory failure. | / | Renal: Need for renal replacement therapy;  Brain: West Haven grade 3–4 HE;  Respiratory: Need for non-invasive or mechanical ventilation;  Circulatory: Shock requiring vasopressor support. |
| ACLF diagnostic model | ACLF is divided into 3 grades of increasing severity.  Grade 1 includes 3 subgroups: (1) single kidney failure; (2) single liver, coagulation, circulation or respiration failure with either kidney dysfunction, brain dysfunction, or both; (3) single brain failure and kidney dysfunction; Grade 2: 2 organ failures; Grade 3: 3 or more organ failures. | ACLF is divided into 3 grades of increasing severity.  Grade 1 includes 4 subgroups: (1) single liver failure and either INR ≥1.5, kidney dysfunction, brain dysfunction, or any combination of these alterations; (2) single kidney failure; (3) single coagulation, circulation or respiration failure with either kidney dysfunction, brain dysfunction, or both; (4) brain failure alone plus kidney dysfunction; Grade 2: 2 organ failures; Grade 3: 3 or more organ failures. | ACLF is classified into 4 grades of increasing severity.  Grade 1 includes patients with a single organ failure involving the liver, kidney, coagulation, or respiration, as well as patients with single brain or circulatory failure accompanied by at least one additional organ dysfunction (subscore = 2) in another organ system; Grade 2: 2 organ failures; Grade 3: 3 or more organ failures; Grade 4: 4 or more organ failures. | TB ≥ 5 mg/dl and INR ≥ 1.5 or prothrombin activity ≤ 40%) accompanied by ascites and/or hepatic encephalopathy within 4 weeks. | Two or more extrahepatic organ failures. |
| Prognostic score | CLIF-OF score  CLIF-C ACLF score | COSSH-ACLF score  COSSH-ACLF II score | A-TANGO OF score  ATANGO-ACLF CRP score  ATANGO-ACLF WBC score | AARC score | NACSELD-ACLF score |

AARC, the APASL ACLF Research Consortium; ACLF, acute-on-chronic liver failure; A-TANGO ACLF-CRP, A-TANGO ACLF-C reactive protein; A-TANGO ACLF-WBC, A-TANGO ACLF-white blood cell; AKI, acute kidney injury; A-TANGO OF, A-TANGO organ failure; CLIF-C ACLF, Chronic Liver Failure Consortium ACLF; CLIF-OF, Chronic Liver Failure-organ failure; COSSH-ACLF, Chinese Group on the Study of Severe Hepatitis B-ACLF; COSSH-ACLF II, Chinese Group on the Study of Severe Hepatitis B-ACLF II; FiO2, fraction of inspired oxygen; HBV-SOFA, Hepatitis B Virus-Related Sequential Organ Failure Assessment; HE, hepatic encephalopathy; INR, international normalised ratio; MAP, mean arterial pressure; NACSELD-ACLF, North American Consortium for the Study of End-Stage Liver Disease-ACLF; PaO2, partial pressure of arterial oxygen; RRT, renal replacement therapy; SpO2, pulse oximetric saturation; TB, total bilirubin.

**Supplemental Table 2.** Calculation formula of prognostic scores.

| Score | Formula |
| --- | --- |
| A-TANGO-ACLF CRP | = 10 * (0.30219 * A-TANGO OF Score + 0.03351 * Age + 0.29446 * log (CRP)) - 9 |
| A-TANGO-ACLF WBC | = 10 * (0.30205 * A-TANGO OF Score + 0.03313 * Age + 0.45920 * log (WBC)) - 8 |
| COSSH-ACLF | = 0.741 * INR + 0.523 * HBV-SOFA + 0.026 * Age + 0.003 * TB (μmol/L) |
| HBV-SOFA | = Kidney score + Brain score + Circulation score + Respiration score |
| COSSH-ACLF II | =1.649 * ln (INR) + 0.457 * HE score (HE grades: 0/1, 1–2/2 and 3–4/3) + 0.425 * ln (Neutrophil) (10^9^/L) + 0.396 * ln (TB) (umol/L) + 0.576 * ln (Urea) + 0.033 * Age |
| CLIF-C ACLF | =10 * (0.33 * CLIF-OFs + 0.04 * Age + 0.63 * ln (WBC) - 2) |
| AARC | = TB score + HE score + INR score + Lactate score + Creatinine score |
| NACSELD-ACLF | = NACSELD-ACLF grade |
| A-TANGO OF | = Liver score + Kidney score + Brain score + Coagulation score + Circulation score + Respiration score |
| CLIF-OF | = Liver score + Kidney score + Brain score + Coagulation score + Circulation score + Respiration score |
| MELD | = 9.57 * ln (Creatinine)(mg/dl) + 3.78 * ln (TB)(mg/dl) + 11.2 * ln (INR) + 6.43 * Etiologies |
| MELD-Na | = MELD score - Na - (0.025 * MELD score * (140 - Na)) +140 |

AARC, the APASL ACLF Research Consortium; ACLF, acute-on-chronic liver failure; A-TANGO ACLF-CRP, A-TANGO ACLF-C reactive protein; A-TANGO ACLF-WBC, A-TANGO ACLF-white blood cell; A-TANGO OF, A-TANGO organ failure; CLIF-C ACLF, Chronic Liver Failure Consortium ACLF; CLIF-OF, Chronic Liver Failure-organ failure; COSSH-ACLF, Chinese Group on the Study of Severe Hepatitis B-ACLF; COSSH-ACLF II, Chinese Group on the Study of Severe Hepatitis B-ACLF II; HBV-SOFA, Hepatitis B Virus-Related Sequential Organ Failure Assessment; HE, hepatic encephalopathy; INR, international normalised ratio; MELD, Model for End-Stage Liver Disease; MELD-Na, Model for End-Stage Liver Disease-sodium; NACSELD-ACLF, North American Consortium for the Study of End-Stage Liver Disease-ACLF; TB, total bilirubin.

**Supplemental Table 3.** Comprehensive comparison of five ACLF diagnostic models based on classification agreement.

| Diagnostic models | NACSELD-ACLF | EASL-CLIF | APASL-ACLF | A-TANGO | COSSH-ACLF |
| --- | --- | --- | --- | --- | --- |
| NACSELD-ACLF | Reference | - | - | - | - |
| EASL-CLIF | <0.001 | Reference | - | - | - |
| APASL-ACLF | <0.001 | <0.001 | Reference | - | - |
| A-TANGO | <0.001 | <0.001 | <0.001 | Reference | - |
| COSSH-ACLF | <0.001 | <0.001 | <0.001 | 0.43 | Reference |

McNemar's test P-values indicate pairwise comparisons between diagnostic models. ACLF, acute-on-chronic liver failure; APASL, Asian Pacific Association for the Study of the Liver; COSSH, Chinese Group on the Study of Severe Hepatitis B; EASL-CLIF, European Association for the Study of the Liver-Chronic Liver Failure Consortium; NACSELD, North American Consortium for the Study of End-Stage Liver Disease.

**Supplemental Table 4.** Characteristics of no ACLF patients according to five diagnostic models.

| Characteristic | No A-TANGO  (n=2236) | No EASL-CLIF  (n=2664) | No COSSH-ACLF  (n=2215) | No APASL-ACLF  (n=2933) | No NACSELD-ACLF  (n=3277) | P value |
| --- | --- | --- | --- | --- | --- | --- |
| Age (years) | 56 ± 11 | 56 ± 12 | 56 ± 11 | 55 ± 12 | 55 ± 12 | <0.001 |
| Male (no.) | 67.8% (1517) | 69.4% (1849) | 67.2% (1489) | 69.8% (2047) | 70.5% (2309) | 0.063 |
| Complication (no.) |  |  |  |  |  |  |
| Hepatic encephalopathy | 8.1% (182) | 7.6% (203) | 9.1% (201) | 13.3% (391) | 14.5% (475) | <0.001 |
| Gastrointestinal bleeding | 35.6% (797) | 32.6% (868) | 37.4% (828) | 31.3% (914) | 28.6% (936) | <0.001 |
| Ascites | 65.6% (1467) | 66.6% (1773) | 64.7% (1434) | 65.5% (1921) | 68.9% (2258) | 0.0084 |
| Infection | 53.9% (1206) | 56.3% (1500) | 52.8% (1170) | 57.3% (1681) | 58.2% (1907) | <0.001 |
| Etiologies (no.) |  |  |  |  |  |  |
| HBV related | 48.7% (1090) | 50.2% (1337) | 48.7% (1079) | 49.5% (1451) | 51.8% (1697) | 0.12 |
| Alcohol related | 25.4% (568) | 25.7% (685) | 24.8% (549) | 26.9% (789) | 26.0% (851) | 0.51 |
| Autoimmune related | 11.4% (255) | 10.7% (286) | 11.0% (244) | 10.5% (308) | 10.0% (328) | 0.53 |
| Others | 19.5% (437) | 18.7% (497) | 20.0% (443) | 18.7% (548) | 17.8% (582) | 0.26 |
| Laboratory data |  |  |  |  |  |  |
| ALT (U/L) | 27.0 (17.0, 52.0) | 30.0 (18.0, 65.0) | 26.0 (17.0, 47.0) | 31.0 (18.0, 70.0) | 33.0 (19.0, 83.0) | <0.001 |
| AST (U/L) | 42.0 (27.0, 83.0) | 48.0 (28.7, 100.0) | 40.0 (26.0, 73.0) | 50.0 (29.0, 107.0) | 55.2 (31.0, 117.0) | <0.001 |
| ALP (U/L) | 108.0 (73.6, 150.0) | 111.0 (77.0, 154.0) | 102.0 (72.0, 146.0) | 111.0 (77.0, 155.0) | 114.0 (80.0, 156.0) | <0.001 |
| ALB (g/L) | 29.9 ± 5.3 | 29.8 ± 5.3 | 29.9 ± 5.3 | 29.8 ± 5.2 | 29.7 ± 5.2 | 0.80 |
| TB (μmol/L) | 37.4 (18.7, 109.3) | 49.9 (20.6, 162.2) | 34.7 (18.1, 94.8) | 58.2 (21.2, 221.0) | 85.1 (23.9, 267.2) | <0.001 |
| TG (mmol/L) | 0.9 (0.7, 1.2) | 0.9 (0.7, 1.3) | 0.9 (0.6, 1.2) | 0.9 (0.7, 1.3) | 0.9 (0.7, 1.3) | <0.001 |
| TCH (mmol/L) | 2.7 (2.1, 3.4) | 2.6 (2.0, 3.2) | 2.7 (2.1, 3.4) | 2.5 (1.9, 3.2) | 2.4 (1.8, 3.2) | <0.001 |
| Creatinine (μmol/L) | 69.0 (57.0, 84.0) | 69.0 (57.0, 83.0) | 69.0 (57.0, 83.3) | 71.0 (58.0, 89.0) | 71.0 (57.0, 88.9) | <0.001 |
| Serum urea (mmol/L) | 5.5 (4.0, 7.8) | 5.5 (4.0, 7.8) | 5.6 (4.0, 7.9) | 5.8 (4.0, 8.7) | 5.7 (4.0, 8.5) | <0.001 |
| Sodium (mmol/L) | 139.0 (136.0, 142.0) | 139.0 (136.0, 141.0) | 137.0 (136.6, 137.0) | 137.0 (135.0, 137.0) | 138.1 (135.0, 141.0) | <0.001 |
| C-reactive protein (mg/L) | 7.8 (3.2, 17.6) | 8.9 (3.3, 19.4) | 7.3 (3.2, 17.2) | 9.2 (3.4, 20.0) | 9.9 (3.8, 20.7) | <0.001 |
| White blood cell count (10^9^/L) | 4.2 (2.8, 6.4) | 4.5 (3.0, 6.9) | 4.1 (2.8, 6.3) | 4.7 (3.1, 7.3) | 4.9 (3.2, 7.7) | <0.001 |
| Neutrophil count (10^9^/L) | 2.6 (1.7, 4.2) | 2.9 (1.8, 4.8) | 2.5 (1.6, 4.2) | 3.0 (1.8, 5.3) | 3.2 (1.9, 5.6) | <0.001 |
| Haemoglobin (g/L) | 95.4 ± 25.6 | 96.6 ± 26.4 | 94.1 ± 26.0 | 96.0 ± 26.7 | 97.4 ± 26.7 | <0.001 |
| Haematocrit (%) | 28.5 ± 7.0 | 28.7 ± 7.1 | 28.2 ± 7.1 | 28.4 ± 7.3 | 28.7 ± 7.3 | 0.063 |
| Platelet count (10^9^/L) | 70.0 (46.0, 108.0) | 71.0 (46.0, 110.0) | 69.0 (45.0, 107.3) | 70.0 (45.0, 111.0) | 71.0 (45.0, 111.0) | 0.63 |
| INR | 1.4 (1.2, 1.6) | 1.4 (1.3, 1.7) | 1.4 (1.2, 1.6) | 1.5 (1.3, 1.8) | 1.5 (1.3, 1.9) | <0.001 |
| Scores |  |  |  |  |  |  |
| A-TANGO ACLF-CRP | 32.9 ± 5.0 | 34.0 ± 5.6 | 33.1 ± 5.3 | 35.1 ± 6.7 | 35.5 ± 6.6 | <0.001 |
| A-TANGO ACLF-WBC | 34.0 ± 4.7 | 35.1 ± 5.3 | 34.3 ± 5.0 | 36.3 ± 6.5 | 36.9 ± 6.6 | <0.001 |
| COSSH-ACLF | 5.0 ± 0.5 | 5.2 ± 0.7 | 5.0 ± 0.6 | 5.4 ± 1.2 | 5.5 ± 1.2 | <0.001 |
| COSSH-ACLF II | 5.9 ± 0.9 | 6.1 ± 1.0 | 5.8 ± 1.0 | 6.3 ± 1.3 | 6.4 ± 1.3 | <0.001 |
| CLIF-C ACLF | 33.6 ± 6.8 | 34.8 ± 7.3 | 33.6 ± 6.9 | 36.4 ± 8.7 | 37.1 ± 8.7 | <0.001 |
| A-TANGO OF | 7.0 (6.0, 7.0) | 7.0 (6.0, 8.0) | 7.0 (6.0, 7.0) | 7.0 (6.0, 9.0) | 7.0 (6.0, 9.0) | <0.001 |
| CLIF-OF | 6.0 (6.0, 7.0) | 7.0 (6.0, 8.0) | 6.0 (6.0, 7.0) | 7.0 (6.0, 8.0) | 7.0 (6.0, 9.0) | <0.001 |
| MELD-Na | 11.7 ± 7.0 | 13.2 ± 7.7 | 11.3 ± 6.7 | 15.1 ± 9.5 | 16.1 ± 9.6 | <0.001 |
| Transplant-free mortality (no.) |  |  |  |  |  |  |
| 28 days | 3.8% (79) | 6.2% (151) | 3.7% (75) | 10.4% (280) | 11.7% (345) | <0.001 |
| 90 days | 8.1% (157) | 12.3% (282) | 7.4% (140) | 17.9% (449) | 19.7% (540) | <0.001 |

Among 3,370 hospitalized patients with acutely decompensated cirrhosis, no ACLF patients were identified according to the five diagnostic models. P values represent differences in clinical characteristics among no ACLF groups defined by the five diagnostic models. ACLF, acute-on-chronic liver failure; ALB, albumin; ALP, alkaline phosphatase; ALT, alanine aminotransferase; AST, aspartate aminotransferase; APASL, Asian Pacific Association for the Study of the Liver; CLIF-C, Chronic Liver Failure Consortium; COSSH, Chinese Group on the Study of Severe Hepatitis B; EASL-CLIF, European Association for the Study of the Liver-Chronic Liver Failure Consortium; HBV, hepatitis B virus; INR, international normalised ratio; MELD-Na, model for end-stage liver disease-sodium; NACSELD, North American Consortium for the Study of End-Stage Liver Disease; OF, organ failure; TB, total bilirubin; TCH, total cholesterol; TG, triglycerides.

**Supplemental Table 5.** Comparison of concordance index among various prognostic scores for predicting 28–day liver transplant-free mortality among patients with AARC data.

| Score | CLIF-OF | A-TANGO OF | COSSH-ACLF | COSSH-ACLF II | AARC | NACSELD-ACLF | MELD-Na |
| --- | --- | --- | --- | --- | --- | --- | --- |
| CLIF-OF | 0.771 (0.721–0.821) | – | – | – | – | – | – |
| A-TANGO OF | 0.17 | 0.793 (0.748–0.838) | – | – | – | – | – |
| COSSH-ACLF | 0.19 | 0.78 | 0.789 (0.742–0.836) | – | – | – | – |
| COSSH-ACLF II | 0.51 | 0.79 | 0.91 | 0.787 (0.738–0.835) | – | – | – |
| AARC | 0.82 | 0.18 | 0.22 | 0.33 | 0.766 (0.719–0.813) | – | – |
| NACSELD-ACLF | <0.001 | <0.001 | <0.001 | <0.001 | <0.001 | 0.571 (0.528–0.613) | – |
| MELD-Na | 0.13 | 0.017 | 0.019 | 0.018 | 0.15 | <0.001 | 0.733 (0.678–0.787) |

Values on the diagonal represent C-index with 95% confidence intervals; P values below the diagonal represent p-values for pairwise comparisons. AARC, the APASL ACLF Research Consortium; ACLF, acute-on-chronic liver failure; CLIF-OF, Chronic Liver Failure-organ failure; C-index, concordance index; COSSH-ACLF, Chinese Group on the Study of Severe Hepatitis B-ACLF; COSSH-ACLF II, Chinese Group on the Study of Severe Hepatitis B-ACLF II; IDI, Integrated Discrimination Improvement; MELD-Na, Model for End-Stage Liver Disease-sodium; NACSELD-ACLF, North American Consortium for the Study of End-Stage Liver Disease-ACLF.

**Supplemental Table 6.** Comparison of concordance index among various prognostic scores for predicting 28–day liver transplant-free mortality according to the A-TANGO diagnostic model.

| Score | A-TANGO ACLF-CRP | A-TANGO ACLF-WBC | COSSH-ACLF | COSSH-ACLF II | CLIF-C ACLF | NACSELD-ACLF | A-TANGO OF | CLIF-OF | MELD-Na |
| --- | --- | --- | --- | --- | --- | --- | --- | --- | --- |
| A-TANGO ACLF-CRP | 0.703 (0.666–0.740) | – | – | – | – | – | – | – | – |
| A-TANGO ACLF-WBC | 0.0015 | 0.719 (0.683–0.754) | – | – | – | – | – | – | – |
| COSSH-ACLF | 0.0033 | 0.044 | 0.745 (0.712–0.778) | – | – | – | – | – | – |
| COSSH-ACLF II | <0.001 | 0.0047 | 0.33 | 0.758 (0.725–0.791) | – | – | – | – | – |
| CLIF-C ACLF | 0.25 | 0.98 | 0.042 | <0.001 | 0.718 (0.682–0.754) | – | – | – | – |
| NACSELD-ACLF | <0.001 | <0.001 | <0.001 | <0.001 | <0.001 | 0.543 (0.520–0.566) | – | – | – |
| A-TANGO OF | 0.57 | 0.63 | 0.013 | 0.0096 | 0.71 | <0.001 | 0.712 (0.676–0.747) | – | – |
| CLIF-OF | 0.65 | 0.73 | 0.0063 | 0.0087 | 0.71 | <0.001 | 0.98 | 0.712 (0.676–0.748) | – |
| MELD-Na | 0.38 | 0.11 | <0.001 | <0.001 | 0.095 | <0.001 | 0.15 | 0.14 | 0.683 (0.645–0.722) |

Values on the diagonal represent C-index with 95% confidence intervals; values below the diagonal represent p-values for pairwise comparisons. ACLF, acute-on-chronic liver failure; A-TANGO ACLF-CRP, A-TANGO ACLF-C reactive protein; A-TANGO ACLF-WBC, A-TANGO ACLF-white blood cell; A-TANGO OF, A-TANGO organ failure; CLIF-C ACLF, Chronic Liver Failure Consortium ACLF; CLIF-OF, Chronic Liver Failure-organ failure; C-index, concordance index; COSSH-ACLF, Chinese Group on the Study of Severe Hepatitis B-ACLF; COSSH-ACLF II, Chinese Group on the Study of Severe Hepatitis B-ACLF II; MELD-Na, Model for End-Stage Liver Disease-sodium; NACSELD-ACLF, North American Consortium for the Study of End-Stage Liver Disease-ACLF.

**Supplemental Table 7.** Comparison of concordance index among various prognostic scores for predicting 28–day liver transplant-free mortality according to the EASL-CLIF diagnostic model.

| Score | A-TANGO ACLF-CRP | A-TANGO ACLF-WBC | COSSH-ACLF | COSSH-ACLF II | CLIF-C ACLF | NACSELD-ACLF | A-TANGO OF | CLIF-OF | MELD-Na |
| --- | --- | --- | --- | --- | --- | --- | --- | --- | --- |
| A-TANGO ACLF-CRP | 0.679 (0.636–0.723) | – | – | – | – | – | – | – | – |
| A-TANGO ACLF-WBC | <0.001 | 0.697 (0.654–0.740) | – | – | – | – | – | – | – |
| COSSH-ACLF | 0.0020 | 0.029 | 0.732 (0.694–0.771) | – | – | – | – | – | – |
| COSSH-ACLF II | 0.0022 | 0.022 | 0.79 | 0.737 (0.698–0.776) | – | – | – | – | – |
| CLIF-C ACLF | 0.085 | 0.59 | 0.080 | 0.018 | 0.703 (0.661–0.746) | – | – | – | – |
| NACSELD-ACLF | <0.001 | <0.001 | <0.001 | <0.001 | <0.001 | 0.546 (0.516–0.577) | – | – | – |
| A-TANGO OF | 0.14 | 0.69 | 0.057 | 0.11 | 1.0 | <0.001 | 0.703 (0.662–0.745) | – | – |
| CLIF-OF | 0.44 | 0.99 | 0.015 | 0.074 | 0.75 | <0.001 | 0.67 | 0.697 (0.656–0.738) | – |
| MELD-Na | 0.17 | 0.038 | <0.001 | <0.001 | 0.021 | <0.001 | 0.012 | 0.041 | 0.643 (0.598–0.688) |

Values on the diagonal represent C-index with 95% confidence intervals; values below the diagonal represent p-values for pairwise comparisons. ACLF, acute-on-chronic liver failure; A-TANGO ACLF-CRP, A-TANGO ACLF-C reactive protein; A-TANGO ACLF-WBC, A-TANGO ACLF-white blood cell; A-TANGO OF, A-TANGO organ failure; CLIF-C ACLF, Chronic Liver Failure Consortium ACLF; CLIF-OF, Chronic Liver Failure-organ failure; C-index, concordance index; COSSH-ACLF, Chinese Group on the Study of Severe Hepatitis B-ACLF; COSSH-ACLF II, Chinese Group on the Study of Severe Hepatitis B-ACLF II; EASL-CLIF, European Association for the Study of the Liver-Chronic Liver Failure Consortium; MELD-Na, Model for End-Stage Liver Disease-sodium; NACSELD-ACLF, North American Consortium for the Study of End-Stage Liver Disease-ACLF.

**Supplemental Table 8.** Comparison of concordance index among various prognostic scores for predicting 28–day liver transplant-free mortality according to the COSSH-ACLF diagnostic model.

| Score | A-TANGO ACLF-CRP | A-TANGO ACLF-WBC | COSSH-ACLF | COSSH-ACLF II | CLIF-C ACLF | NACSELD-ACLF | A-TANGO OF | CLIF-OF | MELD-Na |
| --- | --- | --- | --- | --- | --- | --- | --- | --- | --- |
| A-TANGO ACLF-CRP | 0.709 (0.673–0.746) | – | – | – | – | – | – | – | – |
| A-TANGO ACLF-WBC | 0.039 | 0.718 (0.682–0.754) | – | – | – | – | – | – | – |
| COSSH-ACLF | 0.0021 | 0.0080 | 0.749 (0.716–0.782) | – | – | – | – | – | – |
| COSSH-ACLF II | 0.0022 | 0.0054 | 0.62 | 0.756 (0.722–0.789) | – | – | – | – | – |
| CLIF-C ACLF | 0.71 | 0.69 | 0.0093 | <0.001 | 0.714 (0.678–0.751) | – | – | – | – |
| NACSELD-ACLF | <0.001 | <0.001 | <0.001 | <0.001 | <0.001 | 0.543 (0.520–0.566) | – | – | – |
| A-TANGO OF | 0.57 | 0.93 | 0.0054 | 0.023 | 0.86 | <0.001 | 0.717 (0.682–0.753) | – | – |
| CLIF-OF | 0.97 | 0.58 | <0.001 | 0.0066 | 0.74 | <0.001 | 0.48 | 0.709 (0.673–0.745) | – |
| MELD-Na | 0.093 | 0.031 | <0.001 | <0.001 | 0.045 | <0.001 | 0.020 | 0.064 | 0.672 (0.633–0.710) |

Values on the diagonal represent C-index with 95% confidence intervals; values below the diagonal represent p-values for pairwise comparisons. ACLF, acute-on-chronic liver failure; A-TANGO ACLF-CRP, A-TANGO ACLF-C reactive protein; A-TANGO ACLF-WBC, A-TANGO ACLF-white blood cell; A-TANGO OF, A-TANGO organ failure; CLIF-C ACLF, Chronic Liver Failure Consortium ACLF; CLIF-OF, Chronic Liver Failure-organ failure; C-index, concordance index; COSSH-ACLF, Chinese Group on the Study of Severe Hepatitis B-ACLF; COSSH-ACLF II, Chinese Group on the Study of Severe Hepatitis B-ACLF II; MELD-Na, Model for End-Stage Liver Disease-sodium; NACSELD-ACLF, North American Consortium for the Study of End-Stage Liver Disease-ACLF.

**Supplemental Table 9.** Comparison of concordance index among various prognostic scores for predicting 28–day liver transplant-free mortality according to the APASL-ACLF diagnostic model.

| Score | A-TANGO-ACLF CRP | A-TANGO ACLF-WBC | COSSH-ACLF | COSSH-ACLF II | CLIF-C ACLF | NACSELD-ACLF | A-TANGO OF | CLIF-OF | MELD-Na |
| --- | --- | --- | --- | --- | --- | --- | --- | --- | --- |
| A-TANGO ACLF-CRP | 0.779 (0.726–0.832) | – | – | – | – | – | – | – | – |
| A-TANGO ACLF-WBC | 0.0027 | 0.800 (0.753–0.848) | – | – | – | – | – | – | – |
| COSSH-ACLF | 0.037 | 0.19 | 0.823 (0.776–0.871) | – | – | – | – | – | – |
| COSSH-ACLF II | 0.039 | 0.20 | 0.95 | 0.822 (0.778–0.867) | – | – | – | – | – |
| CLIF-C ACLF | 0.090 | 0.52 | 0.42 | 0.25 | 0.809 (0.761–0.858) | – | – | – | – |
| NACSELD-ACLF | <0.001 | <0.001 | <0.001 | <0.001 | <0.001 | 0.563 (0.522–0.603) | – | – | – |
| A-TANGO OF | 0.33 | 0.96 | 0.093 | 0.36 | 0.74 | <0.001 | 0.801 (0.751–0.852) | – | – |
| CLIF-OF | 0.39 | 0.91 | 0.096 | 0.39 | 0.78 | <0.001 | 0.91 | 0.803 (0.750–0.856) | – |
| MELD-Na | 0.56 | 0.15 | 0.013 | 0.0084 | 0.070 | <0.001 | 0.14 | 0.12 | 0.761 (0.705–0.817) |

Values on the diagonal represent C-index with 95% confidence intervals; values below the diagonal represent p-values for pairwise comparisons. ACLF, acute-on-chronic liver failure; APASL, The Asian Pacific Association for the Study of the Liver; A-TANGO ACLF-CRP, A-TANGO ACLF-C reactive protein; A-TANGO ACLF-WBC, A-TANGO ACLF-white blood cell; A-TANGO OF, A-TANGO organ failure; CLIF-C ACLF, Chronic Liver Failure Consortium ACLF; CLIF-OF, Chronic Liver Failure-organ failure; C-index, concordance index; COSSH-ACLF, Chinese Group on the Study of Severe Hepatitis B-ACLF; COSSH-ACLF II, Chinese Group on the Study of Severe Hepatitis B-ACLF II; MELD-Na, Model for End-Stage Liver Disease-sodium; NACSELD-ACLF, North American Consortium for the Study of End-Stage Liver Disease-ACLF.

**Supplemental Table 10.** Comparison of concordance index among various prognostic scores for predicting 28–day liver transplant-free mortality according to the NACSELD-ACLF diagnostic model.

| Score | A-TANGO ACLF-CRP | A-TANGO ACLF-WBC | COSSH-ACLF | COSSH-ACLF II | CLIF-C ACLF | NACSELD-ACLF | A-TANGO OF | CLIF-OF | MELD-Na |
| --- | --- | --- | --- | --- | --- | --- | --- | --- | --- |
| A-TANGO ACLF-CRP | 0.722 (0.613–0.830) | – | – | – | – | – | – | – | – |
| A-TANGO ACLF-WBC | 0.17 | 0.748 (0.640–0.855) | – | – | – | – | – | – | – |
| COSSH-ACLF | 0.75 | 0.73 | 0.735 (0.622–0.849) | – | – | – | – | – | – |
| COSSH-ACLF II | 0.013 | 0.079 | 0.044 | 0.825 (0.745–0.906) | – | – | – | – | – |
| CLIF-C ACLF | 0.26 | 0.54 | 0.32 | 0.19 | 0.764 (0.656–0.872) | – | – | – | – |
| NACSELD-ACLF | <0.001 | <0.001 | <0.001 | <0.001 | <0.001 | 0.500 (0.500–0.500) | – | – | – |
| A-TANGO OF | 0.48 | 0.057 | 0.13 | 0.0032 | 0.042 | <0.001 | 0.698 (0.590–0.806) | – | – |
| CLIF-OF | 0.68 | 0.24 | 0.097 | 0.021 | 0.046 | 0.0010 | 0.86 | 0.702 (0.583–0.821) | – |
| MELD-Na | 0.95 | 0.55 | 0.77 | 0.0037 | 0.43 | <0.001 | 0.65 | 0.78 | 0.719 (0.623–0.815) |

Values on the diagonal represent C-index with 95% confidence intervals; values below the diagonal represent p-values for pairwise comparisons. ACLF, acute-on-chronic liver failure; A-TANGO ACLF-CRP, A-TANGO ACLF-C reactive protein; A-TANGO ACLF-WBC, A-TANGO ACLF-white blood cell; A-TANGO OF, A-TANGO organ failure; CLIF-C ACLF, Chronic Liver Failure Consortium ACLF; CLIF-OF, Chronic Liver Failure-organ failure; C-index, concordance index; COSSH-ACLF, Chinese Group on the Study of Severe Hepatitis B-ACLF; COSSH-ACLF II, Chinese Group on the Study of Severe Hepatitis B-ACLF II; MELD-Na, Model for End-Stage Liver Disease-sodium; NACSELD-ACLF, North American Consortium for the Study of End-Stage Liver Disease-ACLF.

**Supplemental Table 11.** Integrated discrimination improvement of prognostic scores for 28–day liver transplant-free mortality among patients with available AARC scores.

| Score | CLIF-OF | A-TANGO OF | COSSH-ACLF | COSSH-ACLF II | AARC | NACSELD-ACLF | MELD-Na |
| --- | --- | --- | --- | --- | --- | --- | --- |
| CLIF-OF | Reference | – | – | – | – | – | – |
| A-TANGO OF | 1.45% (-4.27% to 7.16%) | Reference | – | – | – | – | – |
| COSSH-ACLF | 1.41% (-4.02% to 6.95%) | –0.03% (-6.12% to 5.63%) | Reference | – | – | – | – |
| COSSH-ACLF II | 5.75% (-2.62% to 14.61%) | 4.30% (-5.04% to 12.86%) | 4.33% (-2.57% to 11.47%) | Reference | – | – | – |
| AARC | –0.35% (-8.08% to 7.69%) | –1.80% (-10.57% to 6.87%) | –1.76% (-9.82% to 5.98%) | –6.10% (-15.00% to 2.76%) | Reference | – | – |
| NACSELD-ACLF | –19.17% (-28.25% to -10.14%) ^***^ | –20.61% (-30.24% to -11.62%) ^***^ | –20.58% (-30.47% to -10.86%) ^***^ | –24.91% (-35.00% to -14.06%) ^***^ | –18.81% (-27.82% to -9.47%) ^***^ | Reference | – |
| MELD-Na | –6.84% (-15.36% to 2.50%) | –8.28% (-17.72% to 0.48%) | –8.25% (-16.36% to 0.42%) | –12.58% (-21.16% to -5.01%) ^***^ | –6.48% (-14.60% to 1.79%) | 12.33% (2.51% to 22.08%) ^**^ | Reference |

Data are IDI with 95% confidence intervals. Values below the diagonal represent comparisons between scores (P < 0.05 is indicated by *, P < 0.01 by **, and P < 0.001 by ***). AARC, the APASL ACLF Research Consortium; ACLF, acute-on-chronic liver failure; CLIF-OF, Chronic Liver Failure-organ failure; COSSH-ACLF, Chinese Group on the Study of Severe Hepatitis B-ACLF; COSSH-ACLF II, Chinese Group on the Study of Severe Hepatitis B-ACLF II; IDI, Integrated Discrimination Improvement; MELD-Na, Model for End-Stage Liver Disease-sodium; NACSELD-ACLF, North American Consortium for the Study of End-Stage Liver Disease-ACLF.

**Supplemental Table 12.** Integrated discrimination improvement of various prognostic scores for 28–day liver transplant-free mortality prediction according to the A-TANGO model.

| Score | A-TANGO ACLF-CRP | A-TANGO ACLF-WBC | COSSH-ACLF | COSSH-ACLF II | CLIF-C ACLF | NACSELD-ACLF | A-TANGO OF | CLIF-OF | MELD-Na |
| --- | --- | --- | --- | --- | --- | --- | --- | --- | --- |
| A-TANGO ACLF-CRP | Reference | – | – | – | – | – | – | – | – |
| A-TANGO ACLF-WBC | 1.95%  (0.90% to 3.14%) ^**^ | Reference | – | – | – | – | – | – | – |
| COSSH-ACLF | 0.62%  (-8.52% to 8.18%) | –1.34%  (-10.30% to 6.02%) | Reference | – | – | – | – | – | – |
| COSSH-ACLF II | 8.32%  (3.58% to 12.81%) ^***^ | 6.37%  (2.05% to 10.52%) ^**^ | 7.71%  (1.64% to 14.50%) ^*^ | Reference | – | – | – | – | – |
| CLIF-C ACLF | 2.53%  (-0.23% to 5.55%) | 0.57%  (-1.87% to 2.98%) | 1.91%  (-5.42% to 10.80%) | –5.79%  (-9.38% to -2.08%) ^**^ | Reference | – | – | – | – |
| NACSELD-ACLF | –11.00%  (-15.36% to -6.88%) ^***^ | –12.96%  (-17.50% to -8.51%) ^***^ | –11.62%  (-19.61% to -2.46%) ^*^ | –19.32%  (-25.28% to -13.60%) ^***^ | –13.53%  (-18.12% to -9.31%) ^***^ | Reference | – | – | – |
| A-TANGO OF | 1.05%  (-2.48% to 4.68%) | –0.91%  (-4.21% to 2.54%) | 0.43%  (-6.58% to 9.27%) | –7.28%  (-12.52% to -2.01%) ^*^ | –1.48%  (-5.94% to 3.09%) | 12.05%  (7.34% to 16.73%) ^***^ | Reference | – | – |
| CLIF-OF | 1.15%  (-3.39% to 5.85%) | –0.81%  (-5.27% to 3.93%) | 0.53%  (-6.40% to 9.45%) | –7.18%  (-12.63% to -1.95%) ^**^ | –1.38%  (-5.56% to 2.83%) | 12.15%  (8.02% to 16.62%) ^***^ | 0.10%  (-2.94% to 3.18%) | Reference | – |
| MELD-Na | –3.14%  (-8.50% to 1.87%) | –5.10%  (-10.36% to 0.00%) | –3.76%  (-10.86% to 4.21%) | –11.46%  (-16.26% to -6.95%) ^***^ | –5.67%  (-10.96% to -0.66%) ^*^ | 7.86%  (3.40% to 12.42%) ^***^ | –4.19%  (-9.16% to 0.92%) | –4.29%  (-9.43% to 0.43%) | Reference |

Data are IDI with 95% confidence intervals. Values below the diagonal represent comparisons between scores (P < 0.05 is indicated by *, P < 0.01 by **, and P < 0.001 by ***). ACLF, acute-on-chronic liver failure; A-TANGO ACLF-CRP, A-TANGO ACLF-C reactive protein; A-TANGO ACLF-WBC, A-TANGO ACLF-white blood cell; A-TANGO OF, A-TANGO organ failure; CLIF-C ACLF, Chronic Liver Failure Consortium ACLF; CLIF-OF, Chronic Liver Failure-organ failure; COSSH-ACLF, Chinese Group on the Study of Severe Hepatitis B-ACLF; COSSH-ACLF II, Chinese Group on the Study of Severe Hepatitis B-ACLF II; IDI, Integrated Discrimination Improvement; MELD-Na, Model for End-Stage Liver Disease-sodium; NACSELD-ACLF, North American Consortium for the Study of End-Stage Liver Disease-ACLF.

**Supplemental Table 13.** Integrated discrimination improvement of various prognostic scores for 28–day liver transplant-free mortality prediction according to the EASL-CLIF model.

| Score | A-TANGO ACLF-CRP | A-TANGO ACLF-WBC | COSSH-ACLF | COSSH-ACLF II | CLIF-C ACLF | NACSELD-ACLF | A-TANGO OF | CLIF-OF | MELD-Na |
| --- | --- | --- | --- | --- | --- | --- | --- | --- | --- |
| A-TANGO ACLF-CRP | Reference | – | – | – | – | – | – | – | – |
| A-TANGO ACLF-WBC | 2.17%  (0.80% to 3.47%) ^***^ | Reference | – | – | – | – | – | – | – |
| COSSH-ACLF | –0.70%  (-11.05% to 9.62%) | –2.86%  (-13.30% to 7.51%) | Reference | – | – | – | – | – | – |
| COSSH-ACLF II | 8.19%  (2.36% to 14.07%) ^**^ | 6.03%  (0.06% to 11.76%) ^*^ | 8.89%  (-0.15% to 16.02%) | Reference | – | – | – | – | – |
| CLIF-C ACLF | 2.49%  (-0.98% to 5.81%) | 0.32%  (-2.66% to 3.07%) | 3.18%  (-7.52% to 12.45%) | –5.71%  (-10.92% to -0.87%) ^*^ | Reference | – | – | – | – |
| NACSELD-ACLF | –9.99%  (-15.27% to -5.41%) ^***^ | –12.16%  (-17.90% to -7.10%) ^***^ | –9.29%  (-19.96% to 0.06%) | –18.18%  (-25.22% to -10.99%) ^***^ | –12.48%  (-18.00% to -7.17%) ^***^ | Reference | – | – | – |
| A-TANGO OF | 2.98%  (-1.08% to 7.24%) | 0.81%  (-3.25% to 5.10%) | 3.67%  (-6.03% to 13.67%) | –5.22%  (-11.91% to 1.81%) | 0.49%  (-4.94% to 5.82%) | 12.97%  (7.30% to 18.72%) ^***^ | Reference | – | – |
| CLIF-OF | 1.86%  (-3.60% to 7.66%) | –0.31%  (-5.61% to 5.55%) | 2.56%  (-7.33% to 12.21%) | –6.33%  (-13.45% to 0.91%) | –0.63%  (-5.80% to 5.18%) | 11.85%  (6.54% to 17.32%) ^***^ | –1.12%  (-5.18% to 2.89%) | Reference | – |
| MELD-Na | –4.68%  (-10.69% to 1.69%) | –6.85%  (-13.32% to -0.41%) | –3.99%  (-13.81% to 4.24%) | –12.88  (-18.04% to -7.33%) ^***^ | –7.17%  (-13.34% to -0.59%) ^*^ | 5.31%  (0.29% to 11.20%) ^*^ | –7.66%  (-13.69% to -1.32%) ^*^ | –6.55%  (-13.07% to 0.21%) | Reference |

Data are IDI with 95% confidence intervals. Values below the diagonal represent comparisons between scores (P < 0.05 is indicated by *, P < 0.01 by **, and P < 0.001 by ***). ACLF, acute-on-chronic liver failure; A-TANGO ACLF-CRP, A-TANGO ACLF-C reactive protein; A-TANGO ACLF-WBC, A-TANGO ACLF-white blood cell; A-TANGO OF, A-TANGO organ failure; CLIF-C ACLF, Chronic Liver Failure Consortium ACLF; CLIF-OF, Chronic Liver Failure-organ failure; COSSH-ACLF, Chinese Group on the Study of Severe Hepatitis B-ACLF; COSSH-ACLF II, Chinese Group on the Study of Severe Hepatitis B-ACLF II; EASL-CLIF, European Association for the Study of the Liver-Chronic Liver Failure Consortium; IDI, Integrated Discrimination Improvement; MELD-Na, Model for End-Stage Liver Disease-sodium; NACSELD-ACLF, North American Consortium for the Study of End-Stage Liver Disease-ACLF.

**Supplemental Table 14.** Integrated discrimination improvement of various prognostic scores for 28–day liver transplant-free mortality prediction according to the COSSH-ACLF model.

| Score | A-TANGO ACLF-CRP | A-TANGO ACLF-WBC | COSSH-ACLF | COSSH-ACLF II | CLIF-C ACLF | NACSELD-ACLF | ATANGO-OF | CLIF-OF | MELD-Na |
| --- | --- | --- | --- | --- | --- | --- | --- | --- | --- |
| A-TANGO ACLF-CRP | Reference | – | – | – | – | – | – | – | – |
| A-TANGO ACLF-WBC | 1.31%  (0.28% to 2.36%) ^**^ | Reference | – | – | – | – | – | – | – |
| COSSH-ACLF | 0.35%  (-7.61% to 7.26%) | –0.96%  (-9.11% to 5.74%) | Reference | – | – | – | – | – | – |
| COSSH-ACLF II | 6.83%  (2.59% to 11.26%) ^**^ | 5.52%  (1.63% to 9.51%) ^**^ | 6.48%  (0.51% to 12.93%) ^*^ | Reference | – | – | – | – | – |
| CLIF-C ACLF | 0.77%  (-2.09% to 3.71%) | –0.53%  (-2.97% to 1.84%) | 0.42%  (-6.45% to 8.60%) | –6.06%  (-9.85% to -2.51%) ^***^ | Reference | – | – | – | – |
| NACSELD-ACLF | –11.56%  (-15.73% to -7.47%) ^***^ | –12.87%  (-17.37% to -8.62%) ^***^ | –11.91%  (-19.91% to -3.42%) ^**^ | –18.39%  (-24.25% to -12.89%) ^***^ | –12.33%  (-16.76% to -8.01%) ^***^ | Reference | – | – | – |
| A-TANGO OF | 0.97%  (-2.37% to 4.34%) | –0.33%  (-3.37% to 3.01%) | 0.62%  (-6.17% to 8.12%) | –5.86%  (-10.79% to -1.13%) ^*^ | 0.20%  (-4.19% to 4.73%) | 12.53%  (8.09% to 17.14%) ^***^ | Reference | – | – |
| CLIF-OF | –0.04%  (-4.45% to 4.28%) | –1.34%  (-5.63% to 2.72%) | –0.39%  (-7.47% to 7.12%) | –6.87%  (-11.99% to -2.09%) ^*^ | –0.81%  (-4.48% to 3.28%) | 11.52%  (7.14% to 15.76%) ^***^ | –1.01%  (-3.79% to 1.72%) | Reference | – |
| MELD-Na | –5.62%  (-10.08% to -1.08%) ^*^ | –6.92%  (-11.47% to -2.35%) ^**^ | –5.97%  (-13.21% to 0.86%) | –12.45%  (-17.10% to -8.35%) ^***^ | –6.39%  (-11.12% to -1.79%) ^**^ | 5.94%  (1.59% to 10.65%) ^**^ | –6.59%  (-11.24% to -2.15%) ^**^ | –5.58%  (-10.32% to -1.26%) ^*^ | Reference |

Data are IDI with 95% confidence intervals. Values below the diagonal represent comparisons between scores (P < 0.05 is indicated by *, P < 0.01 by **, and P < 0.001 by ***). ACLF, acute-on-chronic liver failure; A-TANGO ACLF-CRP, A-TANGO ACLF-C reactive protein; A-TANGO ACLF-WBC, A-TANGO ACLF-white blood cell; A-TANGO OF, A-TANGO organ failure; CLIF-C ACLF, Chronic Liver Failure Consortium ACLF; CLIF-OF, Chronic Liver Failure-organ failure; COSSH-ACLF, Chinese Group on the Study of Severe Hepatitis B-ACLF; COSSH-ACLF II, Chinese Group on the Study of Severe Hepatitis B-ACLF II; IDI, Integrated Discrimination Improvement; MELD-Na, Model for End-Stage Liver Disease-sodium; NACSELD-ACLF, North American Consortium for the Study of End-Stage Liver Disease-ACLF.

**Supplemental Table 15.** Integrated discrimination improvement of various prognostic scores for 28–day liver transplant-free mortality prediction according to the APASL-ACLF model.

| Score | A-TANGO ACLF-CRP | A-TANGO ACLF-WBC | COSSH-ACLF | COSSH-ACLF II | CLIF-C ACLF | NACSELD-ACLF | ATANGO-OF | CLIF-OF | MELD-Na |
| --- | --- | --- | --- | --- | --- | --- | --- | --- | --- |
| A-TANGO ACLF CRP | Reference | – | – | – | – | – | – | – | – |
| A-TANGO ACLF WBC | 2.94%  (0.95% to 4.94%) ^***^ | Reference | – | – | – | – | – | – | – |
| COSSH-ACLF | 10.27%  (2.93% to 17.27%) ^**^ | 7.33%  (0.31% to 14.14%) ^*^ | Reference | – | – | – | – | – | – |
| COSSH-ACLF II | 10.23%  (2.90% to 17.97%) ^**^ | 7.29%  (0.75% to 13.88%) ^*^ | –0.04%  (-7.97% to 7.48%) | Reference | – | – | – | – | – |
| CLIF-C ACLF | 4.82%  (-0.99% to 10.47%) | 1.88%  (-3.06% to 6.42%) | –5.45%  (-12.72% to 1.93%) | –5.41%  (-10.71% to -0.39%) ^*^ | Reference | – | – | – | – |
| NACSELD-ACLF | –18.02%  (-26.93% to -9.96%) ^***^ | –20.96%  (-30.65% to–12.52%) ^***^ | –28.29%  (-38.77% to -8.45%) ^***^ | –28.25%  (-39.30% to -18.80%) ^***^ | –22.84%  (-31.92% to–14.50%) ^***^ | Reference | – | – | – |
| A-TANGO OF | 2.50%  (-4.08% to 10.13%) | –0.45%  (-6.87% to 6.71%) | –7.77%  (-12.75% to -2.22%) ^**^ | –7.74%  (-16.55% to 0.70%) | –2.32%  (-10.95% to 6.46%) | 20.52%  (12.06% to 30.77%) ^***^ | Reference | – | – |
| CLIF-OF | 6.27%  (-2.65% to 15.96%) | 3.33%  (-5.49% to 12.31%) | –4.00%  (-9.59% to 1.19%) | –3.96%  (-13.68% to 5.17%) | 1.45%  (-7.21% to 9.90%) | 24.29%  (15.22% to 34.65%) ^***^ | 3.77%  (-2.15% to 8.97%) | Reference | – |
| MELD-Na | 0.09%  (-9.00% to 8.49%) | –2.86%  (-11.57% to 5.04%) | –10.18%  (-18.46% to -2.27%) ^*^ | –10.15%  (-18.39% to -2.43%) ^**^ | –4.73%  (-13.31% to 3.63%) | 18.10%  (9.11% to 28.61%) ^***^ | –2.41%  (-10.84% to 4.69%) | –6.18%  (-14.47% to 2.70%) | Reference |

Data are IDI with 95% confidence intervals. Values below the diagonal represent comparisons between scores (P < 0.05 is indicated by *, P < 0.01 by **, and P < 0.001 by ***). ACLF, acute-on-chronic liver failure; APASL, The Asian Pacific Association for the Study of the Liver; A-TANGO ACLF-CRP, A-TANGO ACLF-C reactive protein; A-TANGO ACLF-WBC, A-TANGO ACLF-white blood cell; A-TANGO OF, A-TANGO organ failure; CLIF-C ACLF, Chronic Liver Failure Consortium ACLF; CLIF-OF, Chronic Liver Failure-organ failure; COSSH-ACLF, Chinese Group on the Study of Severe Hepatitis B-ACLF; COSSH-ACLF II, Chinese Group on the Study of Severe Hepatitis B-ACLF II; IDI, Integrated Discrimination Improvement; MELD-Na, Model for End-Stage Liver Disease-sodium; NACSELD-ACLF, North American Consortium for the Study of End-Stage Liver Disease-ACLF

**Supplemental Table 16.** Integrated discrimination improvement of various prognostic scores for 28–day liver transplant-free mortality prediction according to the NACSELD-ACLF model.

| Score | A-TANGO ACLF-CRP | A-TANGO ACLF-WBC | COSSH-ACLF | COSSH-ACLF II | CLIF-C ACLF | NACSELD-ACLF | A-TANGO OF | CLIF-OF | MELD-Na |
| --- | --- | --- | --- | --- | --- | --- | --- | --- | --- |
| A-TANGO ACLF-CRP | Reference | – | – | – | – | – | – | – | – |
| A-TANGO ACLF-WBC | 6.14%  (-0.87% to 14.41%) | Reference | – | – | – | – | – | – | – |
| COSSH-ACLF | 4.65%  (-10.25% to 26.60%) | –1.48%  (-14.41% to 16.77%) | Reference | – | – | – | – | – | – |
| COSSH-ACLF II | 25.47%  (2.43% to 47.87%) ^*^ | 19.33%  (-3.67% to 39.87%) | 20.82%  (-1.38% to 40.23%) | Reference | – | – | – | – | – |
| CLIF-C ACLF | 8.15%  (-7.45% to 24.15%) | 2.01%  (-11.61% to 14.22%) | 3.50%  (-14.49% to 19.29%) | –17.32%  (-37.33% to 2.23%) | Reference | – | – | – | – |
| NACSELD-ACLF | –24.31%  (-53.04% to -4.74%) ^***^ | –30.45%  (-59.88% to -8.56%) ^***^ | –28.96%  (-64.43% to -6.21%) ^***^ | –49.78%  (-80.82% to -24.57%) ^***^ | –32.46%  (-61.82%to -10.00%) ^***^ | Reference | – | – | – |
| A-TANGO OF | –4.45%  (-17.57% to 10.96%) | –10.58%  (-22.49% to 2.17%) | –9.10%  (-21.82% to 0.85%) | –29.91%  (-52.31% to -7.71%) ^**^ | –12.59%  (-29.94% to 5.07%) | 19.86%  (1.64% to 51.92%) ^***^ | Reference | – | – |
| CLIF-OF | –7.04%  (-23.88% to 11.06%) | –13.17%  (-29.19% to 2.85%) | –11.69%  (-23.34% to -2.70%) ^**^ | –32.50%  (-54.35% to -10.37%) ^**^ | –15.18%  (-30.23% to 0.10%) | 17.27%  (1.26% to 47.19%) ^***^ | –2.59%  (-11.78% to 5.78%) | Reference | – |
| MELD-Na | –5.51%  (-28.71% to 13.24%) | –11.65%  (-37.10% to 9.67%) | –10.16%  (-37.06% to 10.86%) | –30.98%  (-55.58% to -10.39%) ^**^ | –13.66%  (-38.48% to 10.09%) | 18.80%  (3.95% to 41.59%) ^***^ | –1.06%  (-23.57% to 17.08%) | 1.53%  (-21.47% to 19.49%) | Reference |

Data are IDI with 95% confidence intervals. Values below the diagonal represent comparisons between scores (P < 0.05 is indicated by *, P < 0.01 by **, and P < 0.001 by ***). ACLF, acute-on-chronic liver failure; A-TANGO ACLF-CRP, A-TANGO ACLF-C reactive protein; A-TANGO ACLF-WBC, A-TANGO ACLF-white blood cell; A-TANGO OF, A-TANGO organ failure; CLIF-C ACLF, Chronic Liver Failure Consortium ACLF; CLIF-OF, Chronic Liver Failure-organ failure; COSSH-ACLF, Chinese Group on the Study of Severe Hepatitis B-ACLF; COSSH-ACLF II, Chinese Group on the Study of Severe Hepatitis B-ACLF II; IDI, Integrated Discrimination Improvement; MELD-Na, Model for End-Stage Liver Disease-sodium; NACSELD-ACLF, North American Consortium for the Study of End-Stage Liver Disease-ACLF.

**Supplemental Table 17.** Clinical characteristics of patients with ACLF defined by the A-TANGO and COSSH-ACLF diagnostic models in the Ambi-Spective cohort from India.

| **Characteristic** | **COSSH-ACLF (n=1697)** | **A-TANGO ACLF (n=1725)** | **P value** |
| --- | --- | --- | --- |
| Age, median (IQR) | 43 (35–52) | 43 (35–52) | 0.70 |
| Male sex, n (%) | 1487 (87.6) | 1514 (87.8) | 0.94 |
| **Precipitating event (n, %)** | | | |
| AH | 698 (41.1) | 723 (41.9) | 1.0 |
| Infection | 193 (11.4) | 204 (11.8) |  |
| AH plus others | 350 (20.6) | 350 (20.3) |  |
| DILI | 128 (7.5) | 125 (7.2) |  |
| Viral hepatitis | 183 (10.8) | 174 (10.1) |  |
| AIH | 12 (0.7) | 12 (0.7) |  |
| UGIB | 13 (0.8) | 13 (0.8) |  |
| Others | 120 (7.1) | 124 (7.2) |  |
| **Etiology (n, %)** | | | |
| AH | 1143 (67.4) | 1178 (68.3) | 1.0 |
| NASH | 99 (5.8) | 103 (6.0) |  |
| Viral hepatitis | 221 (13.0) | 214 (12.4) |  |
| MetALD | 26 (1.5) | 27 (1.6) |  |
| AIH | 89 (5.2) | 90 (5.2) |  |
| AH plus Viral | 81 (4.8) | 77 (4.5) |  |
| NASH plus AIH | 38 (2.2) | 36 (2.1) |  |
| **Complications (n, %)** | | | |
| Ascites | 1637 (96.5) | 1663 (96.4) | 1.0 |
| Hepatic encephalopathy | 994 (58.6) | 981 (56.9) | 0.33 |
| Infection | 1081 (63.7) | 1103 (63.9) | 0.91 |
| **Lab investigations** | | | |
| Haemoglobin (gm/dL) | 9.6 (7.9–11.3) | 9.5 (7.8–11.2) | 0.52 |
| WBC (x10^9^/L) | 12.5 (8.7–18.1) | 12.2 (8.4–18.0) | 0.24 |
| Platelet count (x10^9^/L) | 110.0 (70.0–169.0) | 107.0 (69.0–164.0) | 0.47 |
| INR | 2.2 (1.8–2.7) | 2.2 (1.8–2.7) | 0.19 |
| AST (U/L) | 133.6 (87.0–199.0) | 132.0 (85.0–198.0) | 0.45 |
| ALT (U/L) | 55.0 (35.0–100.0) | 54.0 (34.0–98.0) | 0.44 |
| Bilirubin (mg/dL) | 20.0 (12.6–27.0) | 20.0 (10.1–27.0) | 0.085 |
| Creatinine (mg/dL) | 1.4 (0.8–2.4) | 1.3 (0.8–2.4) | 0.61 |
| Sodium (mmol/L) | 132.0 (128.0–136.0) | 132.0 (128.0–136.0) | 0.77 |
| **Disease Severity scores** | | | |
| MELD-Na score | 32.0 (28.0–36.0) | 30.8 (26.8–35.0) | 0.46 |
| CLIF-C OF score | 11.0 (10.0–13.0) | 11.0 (10.0–12.0) | 0.56 |
| A-TANGO OF score | 11.0 (10.0–13.0) | 11.0 (10.0–13.0) | 0.85 |
| COSSH-ACLF score | 7.2 (6.4–8.1) | 7.1 (6.4–8.1) | 0.63 |
| COSSH-ACLF II score | 9.2 (8.4–10.0) | 9.2 (8.4–10.0) | 0.55 |
| **Transplant-free mortality (n, %)** | | |  |
| 28 days | 847 (49.9) | 852 (49.4) | 0.79 |
| 90 days | 1030 (60.7) | 1037 (60.1) | 0.76 |
| AH, alcoholic hepatitis; AIH, autoimmune hepatitis; ALT, alanine aminotransferase; AST, aspartate aminotransferase; CLIF-C OF, Chronic Liver Failure Consortium Organ Failure; COSSH-ACLF, Chinese Group on the Study of Severe Hepatitis B Acute-on-Chronic Liver Failure; DILI, drug-induced liver injury; INR, international normalized ratio; MELD, Model for End-Stage Liver Disease; MetALD, Metabolic dysfunction-Alcohol associated liver disease; NASH, nonalcoholic steatohepatitis; UGIB, upper gastrointestinal bleeding; WBC, white blood cell count. | | | |

**Supplemental Table 18.** Net reclassification improvement of A-TANGO versus COSSH-ACLF for predicting 28–day liver transplant-free mortality in the Ambi-Spective cohort from India.

| COSSH-ACLF  diagnostic model | A-TANGO diagnostic model (NRI=17.1%) | | | | | |
| --- | --- | --- | --- | --- | --- | --- |
|  | No ACLF | ACLF grade 1 | ACLF grade 2 | ACLF grade 3 | ACLF grade 4 | Total |
| No ACLF | | | | | | |
| Number of patients. n (%) | 233 (65.1) | 112 (31.3) | 13 (3.6) | NA | NA | 358 (17.4) |
| 28–day mortality. n (%) | 38 (16.3) | 21 (18.8) | 0 (0.0) | NA | NA | 59 (16.5) |
| ACLF grade 1 | | | | | | |
| Number of patients. n (%) | 97 (20.8) | 311 (66.7) | 57 (12.2) | 1 (0.2) | NA | 466 (22.7) |
| 28–day mortality. n (%) | 16 (16.5) | 104 (33.4) | 31 (54.4) | 0 (0.0) | NA | 151 (32.4) |
| ACLF grade 2 | | | | | | |
| Number of patients. n (%) | NA | 132 (21.5) | 396 (64.6) | 83 (13.5) | 2 (3.3) | 613 (29.8) |
| 28–day mortality. n (%) | NA | 53 (40.2) | 177 (44.7) | 45 (54.2) | 1 (50.0) | 276 (45.0) |
| ACLF grade 3 | | | | | | |
| Number of patients. n (%) | NA | NA | 86 (13.9) | 305 (49.3) | 227 (36.7) | 618 (30.1) |
| 28–day mortality. n (%) | NA | NA | 39 (45.3) | 205 (67.2) | 176 (77.5) | 420 (68.0) |
| Total | 330 | 555 | 552 | 389 | 229 | 2055 |

ACLF, acute-on-chronic liver failure; COSSH, Chinese Group on the Study of Severe Hepatitis B; LT, liver transplantation; NRI, The Net Reclassification Improvement.

**Supplemental Table 19.** Concordance index among various prognostic scores for predicting 28–day liver transplant-free mortality in the Ambi-Spective cohort from India.

| Score | A-TANGO OF | COSSH-ACLF | COSSH-ACLF II |
| --- | --- | --- | --- |
| A-TANGO OF | 0.706 (0.686–0.725) | – | – |
| COSSH-ACLF | 0.0013 | 0.690 (0.669–0.710) | – |
| COSSH-ACLF II | 0.70 | 0.23 | 0.702 (0.681–0.722) |

Values on the diagonal represent C-index with 95% confidence intervals; values below the diagonal represent p-values for pairwise comparisons. ACLF, acute-on-chronic liver failure; C-index, concordance index; COSSH, Chinese Group on the Study of Severe Hepatitis B; C-index, concordance index; OF, organ failure.

**Supplemental Table 20.** Integrated discrimination improvement of various prognostic scores for 28–day liver transplant-free mortality prediction in the Ambi-Spective cohort from India.

| Score | A-TANGO OF | COSSH-ACLF | COSSH-ACLF II |
| --- | --- | --- | --- |
| A-TANGO OF | Reference | – | – |
| COSSH-ACLF | –4.63% (-5.61% to -3.60%) *** | Reference | – |
| COSSH-ACLF II | –9.07% (-1.12% to -6.96%) *** | –3.61% (-5.23% to -2.01%) *** | Reference |

Data are IDI with 95% confidence intervals. Values below the diagonal represent comparisons between scores (P < 0.05 is indicated by *, P < 0.01 by **, and P < 0.001 by ***). ACLF, acute-on-chronic liver failure; COSSH, Chinese Group on the Study of Severe Hepatitis B; IDI, Integrated Discrimination Improvement; OF, organ failure.

**Supplemental Table 21.** Net reclassification improvement of the A-TANGO^+^/COSSH^+^ diagnostic model compared with the A-TANGO diagnostic model or COSSH diagnostic model only for 28–day liver transplant-free mortality.

| A-TANGO diagnostic model only vs. A-TANGO^+^/COSSH^+^ diagnostic model | | | |
| --- | --- | --- | --- |
| A-TANGO diagnostic model only | A-TANGO^+^/COSSH^+^ diagnostic model (NRI=0.9%) | | |
|  | No ACLF | ACLF | Total |
| No ACLF | | | |
| Number of patients. n (%) | 2067 (100.0) | NA | 2067 (68.7) |
| 28–day mortality. n (%) | 79 (3.8) | NA | 79 (3.8) |
| ACLF | | | |
| Number of patients. n (%) | 157 (16.7) | 783 (83.3) | 940 (31.3) |
| 28–day mortality. n (%) | 17 (10.8) | 288 (36.8) | 305 (32.4) |
| Total | 2224 (74.0) | 783 (26.0) | 3007 |
| COSSH diagnostic model only vs. A-TANGO^+^/COSSH^+^ diagnostic model | | | |
| COSSH diagnostic model only | A-TANGO^+^/COSSH^+^ diagnostic model (NRI=1.2%) | | |
|  | No ACLF | ACLF | Total |
| No ACLF | | | |
| Number of patients. n (%) | 2028 (100.0) | NA | 2028 (67.4) |
| 28–day mortality. n (%) | 75 (3.7) | NA | 75 (3.7) |
| ACLF | | | |
| Number of patients. n (%) | 196 (20.0) | 783 (83.3) | 979 (32.6) |
| 28–day mortality. n (%) | 21 (10.7) | 288 (36.8) | 309 (31.6) |
| Total | 2224 (74.0) | 783 (26.0) | 3007 |

ACLF, acute-on-chronic liver failure; APASL, Asian Pacific Association for the Study of the Liver; COSSH, Chinese Group on the Study of Severe Hepatitis B; EASL-CLIF, European Association for the Study of the Liver-Chronic Liver Failure Consortium; NACSELD, North American Consortium for the Study of End-Stage Liver Disease; NRI, The Net Reclassification Improvement.

**Supplemental Table 22.** Concordance index of various prognostic scores for 28–day liver transplant-free mortality prediction in the ATATGO^+^ /COSSH^+^ group.

| Score | A-TANGO ACLF-CRP | A-TANGO ACLF-WBC | COSSH-ACLF | COSSH-ACLF II | CLIF-C ACLF | NACSELD-ACLF | A-TANGO OF | CLIF-OF | MELD-Na |
| --- | --- | --- | --- | --- | --- | --- | --- | --- | --- |
| A-TANGO ACLF-CRP | 0.696 (0.659–0.734) | – | – | – | – | – | – | – | – |
| A-TANGO ACLF-WBC | 0.028 | 0.707 (0.670–0.744) | – | – | – | – | – | – | – |
| COSSH-ACLF | 0.014 | 0.056 | 0.733 (0.699–0.767) | – | – | – | – | – | – |
| COSSH-ACLF II | 0.0034 | 0.0095 | 0.38 | 0.746 (0.711–0.781) | – | – | – | – | – |
| CLIF-C ACLF | 0.43 | 0.99 | 0.064 | <0.001 | 0.707 (0.669–0.745) | – | – | – | – |
| NACSELD-ACLF | <0.001 | <0.001 | <0.001 | <0.001 | <0.001 | 0.543 (0.518–0.568) | – | – | – |
| A-TANGO OF | 0.75 | 0.30 | 0.0034 | 0.0056 | 0.41 | <0.001 | 0.691 (0.653–0.730) | – | – |
| CLIF-OF | 0.81 | 0.43 | 0.0015 | 0.0051 | 0.39 | <0.001 | 1.0 | 0.692 (0.654–0.729) | – |
| MELD-Na | 0.058 | 0.015 | <0.001 | <0.001 | 0.012 | <0.001 | 0.058 | 0.058 | 0.650 (0.610–0.691) |

Values on the diagonal represent C-index with 95% confidence intervals; P values below the diagonal represent p-values for pairwise comparisons. ACLF, acute-on-chronic liver failure; CLIF-OF, Chronic Liver Failure-organ failure; C-index, concordance index; COSSH-ACLF, Chinese Group on the Study of Severe Hepatitis B-ACLF; COSSH-ACLF II, Chinese Group on the Study of Severe Hepatitis B-ACLF II; MELD-Na, Model for End-Stage Liver Disease-sodium; NACSELD-ACLF, North American Consortium for the Study of End-Stage Liver Disease-ACLF.

**Supplemental Table 23.** Integrated discrimination improvement of various prognostic scores for 28–day liver transplant-free mortality prediction in the ATATGO^+^ /COSSH^+^ group.

| Score | A-TANGO ACLF-CRP | A-TANGO ACLF-WBC | COSSH-ACLF | COSSH-ACLF II | CLIF-C ACLF | NACSELD-ACLF | A-TANGO OF | CLIF-OF | MELD-Na |
| --- | --- | --- | --- | --- | --- | --- | --- | --- | --- |
| A-TANGO ACLF-CRP | Reference | – | – | – | – | – | – | – | – |
| A-TANGO ACLF-WBC | 1.41%  (0.29% to 2.59%) ** | Reference | – | – | – | – | – | – | – |
| COSSH-ACLF | –0.75%  (-9.50% to 6.91%) | –2.16%  (-11.12% to 5.45%) | Reference | – | – | – | – | – | – |
| COSSH-ACLF II | 7.54%  (2.80% to 12.30%) ** | 6.12%  (1.48% to 10.78%) * | 8.29%  (1.56% to 15.35%) ** | Reference | – | – | – | – | – |
| CLIF-C ACLF | 1.86%  (-1.38% to 4.54%) | 0.45%  (-2.20% to 2.69%) | 2.61%  (-5.32% to 11.31%) | –5.68%  (-9.71% to -1.74%) ** | Reference | – | – | – | – |
| NACSELD-ACLF | –10.29%  (-15.05% to -5.90%) *** | –11.70%  (-16.94% to -7.18%) *** | –9.54%  (-18.25% to -0.85%) * | –17.83%  (-24.14% to -11.70%) *** | –12.15%  (-17.31% to -7.45%) *** | Reference | – | – | – |
| A-TANGO OF | –0.14%  (-3.91% to 3.85%) | –1.55%  (-5.19% to 2.14%) | 0.61%  (-7.18% to 9.68%) | –7.67%  (-13.60% to -1.87%) * | –1.99%  (-6.45% to 3.00%) | 10.15%  (5.67% to 15.27%) *** | Reference | – | – |
| CLIF-OF | –0.39%  (-5.22% to 3.89%) | –1.80%  (-6.61% to 2.56%) | 0.36%  (-7.25% to 8.77%) | –7.93%  (-13.50% to -2.59%) ** | –2.25%  (-6.64% to 2.08%) | 9.90%  (5.87% to 14.47%) *** | –0.26%  (-3.54% to 2.64%) | Reference | – |
| MELD-Na | –5.48%  (-11.25% to -0.23%) * | –6.89%  (-12.49% to -1.65%) * | –4.73%  (-12.48% to 2.97%) | –13.02%  (-17.86% to -8.29%) *** | –7.34%  (-12.77% to -1.88%) ** | 4.81%  (0.74% to 9.48%) * | –5.34%  (-10.58% to -0.14%) * | –5.09%  (-9.69% to -0.02%) * | Reference |

Data are IDI with 95% confidence intervals. Values below the diagonal represent comparisons between scores (P < 0.05 is indicated by *, P < 0.01 by **, and P < 0.001 by ***).

ACLF, acute-on-chronic liver failure; A-TANGO ACLF-CRP, A-TANGO ACLF-C reactive protein; A-TANGO ACLF-WBC, A-TANGO ACLF-white blood cell; A-TANGO OF, A-TANGO organ failure; CLIF-C ACLF, Chronic Liver Failure Consortium ACLF; CLIF-OF, Chronic Liver Failure-organ failure; COSSH-ACLF, Chinese Group on the Study of Severe Hepatitis B-ACLF; COSSH-ACLF II, Chinese Group on the Study of Severe Hepatitis B-ACLF II; IDI, Integrated Discrimination Improvement; MELD-Na, Model for End-Stage Liver Disease-sodium; NACSELD-ACLF, North American Consortium for the Study of End-Stage Liver Disease-ACLF.

**Supplemental Table 24.** Sensitivity analyses of multivariable Cox models for 90–day liver transplant-free mortality in the intermediate-risk group.

| Parameter | Model 1 (WBC included)-HR (95% CI) | Model 2 (Neutrophil count instead of WBC)-HR (95% CI) | P value (Model 1) | P value (Model 2) |
| --- | --- | --- | --- | --- |
| Neutrophil count (10^9^/L) | NA | 1.08 (1.01–1.16) | NA | 0.026 |
| Sodium (mmol/L) | 0.96 (0.91–1.01) | 0.96 (0.91–1.01) | 0.12 | 0.11 |
| TCH (mmol/L) | 0.62 (0.43–0.88) | 0.63 (0.44–0.89) | 0.0085 | 0.0092 |
| White blood cell count (10^9^/L) | 1.07 (1.01–1.15) | NA | 0.032 | NA |
| Age (years) | 1.02 (0.99–1.05) | 1.02 (1.00–1.05) | 0.12 | 0.11 |
| Male (yes vs no) | 0.56 (0.31–1.01) | 0.57 (0.31–1.03) | 0.053 | 0.061 |

White blood cell count and neutrophil count were not included simultaneously in the same model because of collinearity; TCH, total cholesterol.

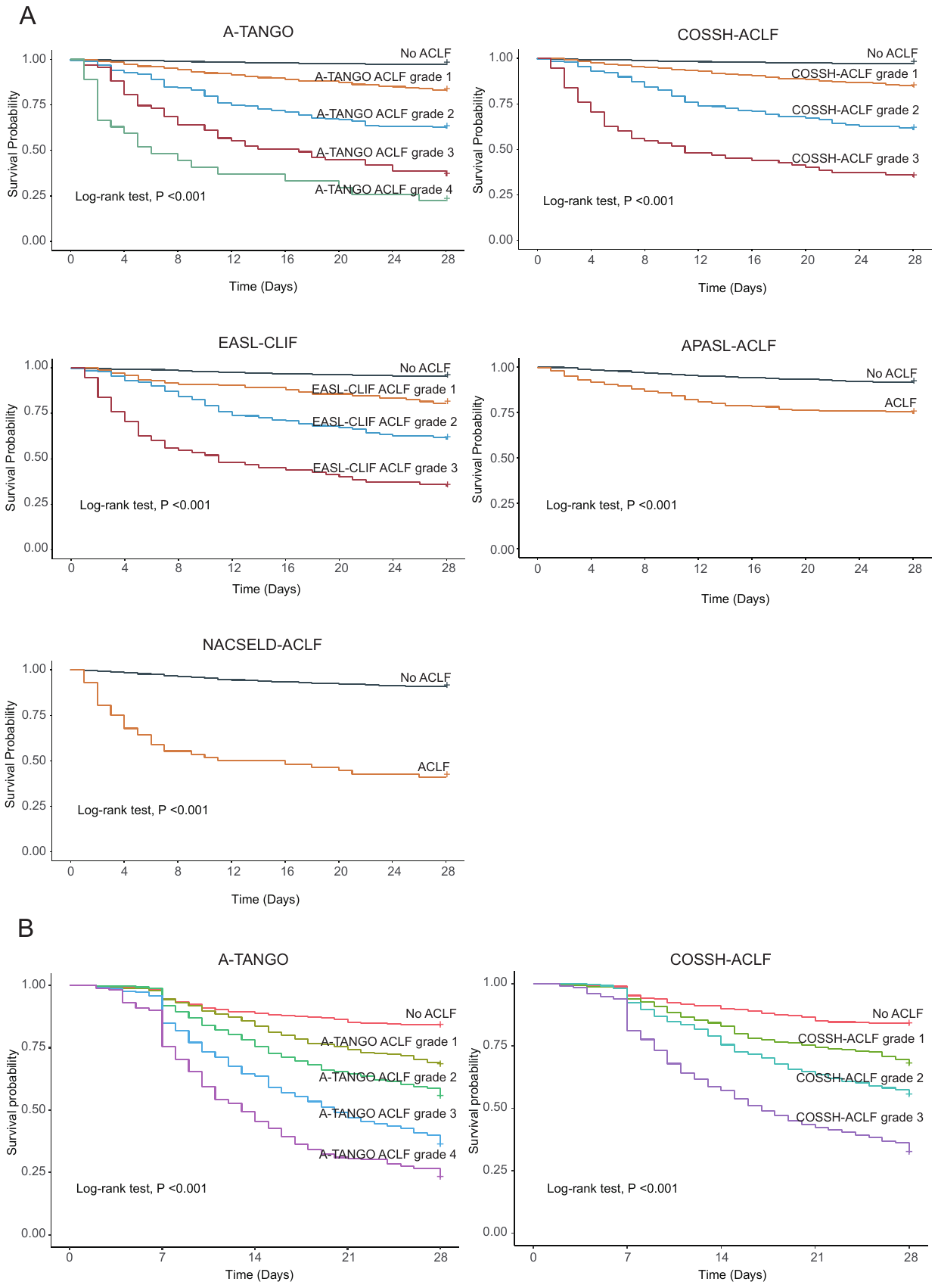
 **Supplemental Figure 1.** Survival stratification according to ACLF grades defined by diferent diagnostic models. (A) Kaplan–Meier curves for 28–day survival stratified by ACLF grades according to five models in the COSSH cohort (n=3,370). (B) Kaplan–Meier curves for 28–day survival stratified by ACLF grades according to A-TANGO and COSSH-ACLF diagnostic models in the Indian Ambi-Spective cohort. ACLF, acute-on-chronic liver failure; APASL, Asian Pacific Association for the Study of the Liver; COSSH, Chinese Group on the Study of Severe Hepatitis B; EASL-CLIF, European Association for the Study of the Liver-Chronic Liver Failure Consortium; NACSELD, North American Consortium for the Study of End-Stage Liver Disease.

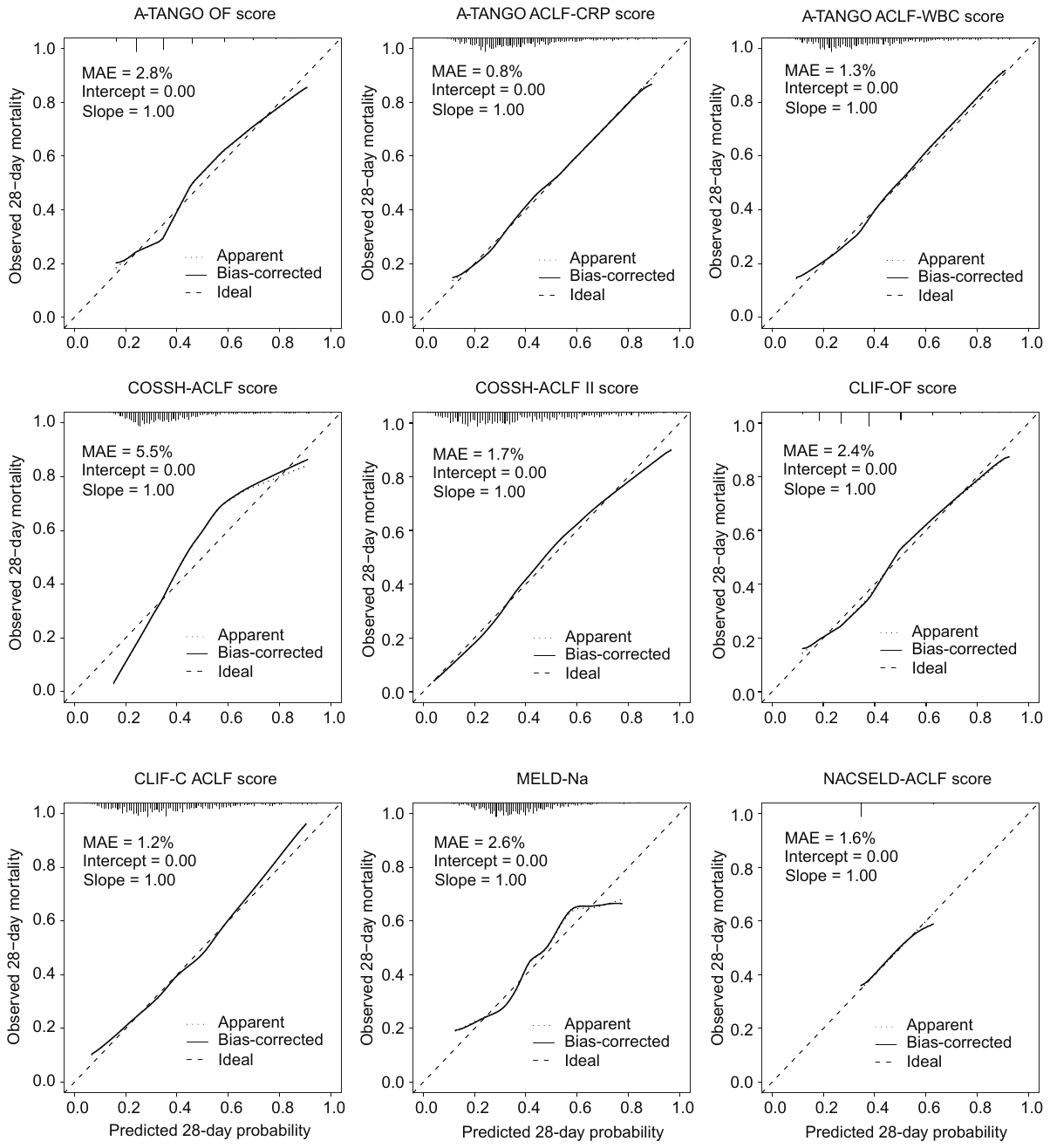

**Supplemental Figure 2.** Calibration of nine prognostic scores for 28–day liver transplant-free mortality in the ATANGO^+^/COSSH^+^ group (n=947). ACLF, acute-on-chronic liver failure; APASL, The Asian Pacific Association for the Study of the Liver; A-TANGO ACLF-CRP, A-TANGO ACLF-C reactive protein; A-TANGO ACLF-WBC, A-TANGO ACLF-white blood cell; A-TANGO OF, A-TANGO organ failure; CLIF-C ACLF, Chronic Liver Failure Consortium ACLF; CLIF-OF, Chronic Liver Failure-organ failure; COSSH-ACLF, Chinese Group on the Study of Severe Hepatitis B-ACLF; COSSH-ACLF II, Chinese Group on the Study of Severe Hepatitis B-ACLF II; MELD-Na, Model for End-Stage Liver Disease-sodium; NACSELD-ACLF, North American Consortium for the Study of End-Stage Liver Disease-ACLF.

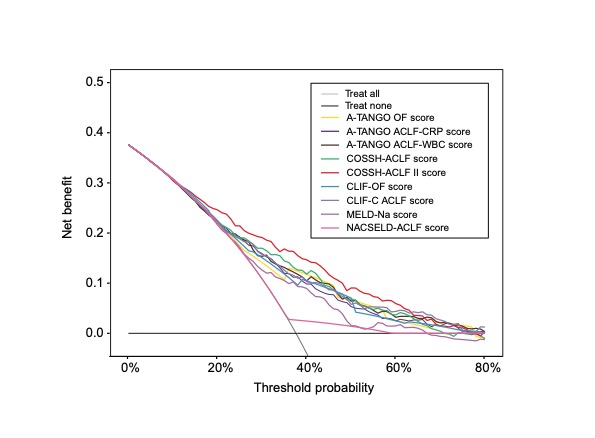

**Supplemental Figure 3.** Decision curve analysis of prognostic scores for 28–day liver transplant-free mortality in the ATANGO^+^/COSSH^+^ group (n=947). ACLF, acute-on-chronic liver failure; CLIF, Chronic Liver Failure; COSSH, Chinese Group on the Study of Severe Hepatitis B; MELD-Na, model for end-stage liver disease-sodium; NACSELD, North American Consortium for the Study of End-Stage Liver Disease; OF, organ failure.
